# Genome-wide pleiotropy analysis identifies novel blood pressure variants and improves its polygenic risk scores

**DOI:** 10.1101/2021.09.08.21263225

**Authors:** Xiaofeng Zhu, Luke Zhu, Heming Wang, Richard S Cooper, Aravinda Chakravarti

**Author notes:** Corresponding author: X. Zhu.

## Abstract

Systolic and diastolic blood pressure (S/DBP) are highly correlated modifiable risk factors for cardiovascular disease (CVD). We report here a bidirectional Mendelian Randomization (MR) and pleiotropy analysis of systolic and diastolic blood pressure (BP) summary statistics from the UKB-ICBP BP genome-wide association study (GWAS) and construct a composite genetic risk score (GRS) by including pleiotropic variants. The composite GRS captures greater (1.11-3.26 fold) heritability for BP traits and increases (1.09- and 2.01-fold) Nagelkerke’s R^2^ for hypertension (HTN) and cardiovascular disease (CVD). We replicated 118 novel BP pleiotropic variants including 18 novel BP loci using summary statistics from the Million Veteran Program (MVP) study. An additional 219 novel BP signals and 40 novel loci were identified after meta-analysis of the UKB-ICBP and MVP summary statistics but without further independent replication. Our study provides further insight into BP regulation and provides a novel way to construct a GRS by including pleiotropic variants for other complex diseases.

## Introduction

Poorly controlled blood pressure (BP) accounts for a large portion of the risk for cardiovascular disease (CVD), stroke, and heart failure^1^. Understanding biological mechanisms for BP regulation could thus potentially help improve BP control and lead to a reduction in the burden of CVD. BP, characterized by systolic and diastolic blood pressure (SBP/DBP), are long-standing risk predictors for CVD. To date, genome-wide association studies (GWAS) have been performed on BP traits by focusing on main effects and in studies that included subjects of diverse ancestry over 1,000 BP-associated loci have been identified^2-14^. Genome-wide search of gene-environment interactions on BP traits have also been recently conducted, however, only a few new gene-environment interactions have been identified, in part owing to low statistical power^13; 14^. Although many GWAS variants are shared between SBP and DBP, both of which are correlated, some seem associated only with SBP or DBP, suggesting evidence of trait-specific biological mechanisms. It has been reported that joint analysis of SBP and DBP leads to the identification of BP variants missed by analyzing SBP or DBP separately^9^. However prior studies have not addressed the mechanisms underlying the SBP-DBP relationship, reflecting arterial stiffness or arterial compliance ^15^. Dissecting the causal relationships of SBP and DBP variants, in particular, whether they affect SBP and DBP through the same (mediation) or different (pleiotropic) paths, and how many pleiotropic variants contribute jointly to these highly correlated traits, is thus important for understanding the biology of BP regulation.

Genetic risk scores (GRS) are constructed as weighted linear combinations of individual variant effects estimated from GWAS to predict individual-level risk of a common disease. An overall GRS is the average of SBP- and DBP-specific GRS^3; 6^. However, published BP GRS’s have explained ∼6% of the heritability of SBP and DBP, and have limited predictive power for HTN and CVD. GWAS of gene-age interaction analysis have also identified genetic variants with age-dependent effect sizes, including for BP^16; 17^, lipid levels^18^ and BMI^19^. A recent study based on a proportional hazards model reported age-varying risk profiles in nine diseases, including HTN^20^. However, these studies were under powered because the interactive contribution by variant and age is often weak.

In this study we address the mechanistic relationship between SBP and DBP by performing a bidirectional Mendelian Randomization (MR) ^21^ and GWAS pleiotropy analysis using summary statistics from >750,000 subjects of European ancestry from the UKB and ICBP consortium^6^, followed by summary statistics of 318,891 multi-ethnic subjects from the Million Veteran Program (MVP) ^12^. We searched for novel BP variants with pleiotropic effects and constructed a composite GRS using variants with and without pleiotropic effects, and studied the age-varying effects of GRS for prediction of BP, HTN and CVD in European, African and Asian descent individuals.

## Material and Methods

### Summary statistics of UK Biobank (UKB) and International Consortium for Blood Pressure (ICBP)

UKB and the ICBP consists of data on 458,577 UK and 299,024 European des cent subjects. GWAS of SBP and DBP were conducted in UKB and ICBP separately and the results were meta-analyzed.^3; 6^ Our analysis was based on the summary results from the UKB and ICBP GWAS that were calculated based on up to 757,601 participants and ∼7.1 M genotyped and imputed SNPs with MAF ≥ 1% for variants present in both the UKB data and ICBP meta-analysis for SBP, DBP and pulse pressure (PP defined as SBP-DBP).

### Summary statistics of the Million Veteran Program (MVP)

The BP summary statistics of the Million Veteran Program (MVP) consists of 318,891 predominantly male multiethnic participants from Hispanic, non-Hispanic whites, blacks, Asians and Native Americans.^12^ There were 18.2M genotyped and imputed SNPs in the summary statistics. The MVP data were used for replication analysis as well as meta-analysis with UKB-ICBP.

### UKB individual level data

Participants in the UKB were genotyped using a custom Affymetrix UK Biobank Axiom array ^22^. Genotypes were imputed by the UKB using the Haplotype Reference Consortium reference panel ^23^; we retained variants with imputation Rsq > 0.3. Related individuals with pairwise kinship coefficient greater than 0.0884 (suggested by UKB) were removed from analysis, resulting in 451,174 individuals of European, African and Asian ancestries. The principal components were calculated by UKB with genotype data within each ancestry to account for population structure.

We analyzed three BP traits in UKB: SBP, DBP and PP. We calculated the mean SBP and DBP values from two baseline BP measurements and added 15 and 10 mmHg to SBP and DBP, respectively, for individuals who took antihypertensive medications. Hypertensive cases were defined as either SBP ≥140 or DBP≥90 or taking antihypertensive medications. CVD cases in UKB were defined using self-reported baseline information and the ICD9 and ICD10 diagnostic codes on hospital admissions. The CVD cases includes ICD9 (“4109”, “4119”,”4129”, “4139”, “4140”, “4141”, “4148”, “4149”) and ICD10 (I210, I211, I212, I213, I214, I219, I21X, I220, I221, I228, I229, I230, I231, I232, I233, 234, I235, I236, I238, I240, I241, I248, I249, I250, I251, I252, I253, I254, I255, I256, I258, I259) codes These data identified 35,968 CVD cases in subjects with European, African and Asian ancestries. The study was approved by the Case Western Reserve University Institutional Review Board (STUDY20180592).

### Mendelian Randomization analysis

We performed a bi-directional MR analysis of SBP and DBP by applying the software IMRP^24^ and MRmix ^25^, as well as estimated the causal contributions of BP on Coronary Artery Disease (CAD), myocardial infarction (MI) and Stroke. Considering an exposure (*Y*_1_) and an outcome (*Y*_2_) the following association model as described in **Figure 1A** was used:

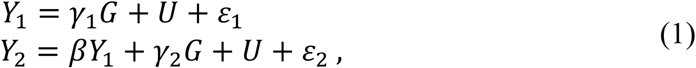

where *G* is a genetic instrumental variable (IV), *γ*_1_ and *γ*_2_ are the direct contributions of *G* to exposure and outcome, β is the causal effect of the exposure *Y*_1_ to the outcome *Y*_2_, U represents confounding factors and ε_1_ and ε_2_ are error terms, respectively. MR analysis estimates the causal effect β through the genetic IV *G*. A valid genetic IV satisfies *γ*_1_ ≠ 0 and *γ*_2_ = 0, representing the genetic contribution to outcome through the mediation of the exposure. We termed these variants (*γ*_2_ = 0) as mediation variants. We define a pleiotropic variant as one with *γ*_1_ ≠ 0 and *γ*_2_ ≠ 0, interpreted as the genetic contributions to exposure and outcome through two independent paths (or a pleiotropic path) (**Figure 1A**). IMRP is an iterative approach combining the pleiotropy test and the MR analysis. The iteration starts by performing MR-Egger analysis^26^ to estimate the causal effect of an exposure to outcome, following by inverse variance weighted (IVW)^27; 28^ analysis until the causal effect estimate converges. The causal effect is estimated by IVW after excluding all identified pleiotropic variants. At each iteration step, IMRP perform pleiotropy test to update which genetic instrument variants show pleiotropy (P<0.05) using the test:

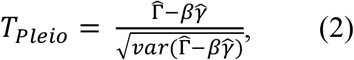

where 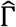 and 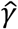 are the estimated effect sizes of a genetic IV on exposure and the outcome, respectively, and *β* is the causal estimate which is updated at each iterative step. We have previously shown that *T*_*Pleio*_ tests the null hypothesis *γ*_2_ = 0 ^24^. In MR analysis, a valid IV satisfies *γ*_1_ ≠ 0, therefore rejecting null hypothesis *γ*_2_ = 0 and suggesting a pleiotropic effect. IMRP takes advantage of MR-Egger, which is less biased, and IVW, which is more efficient. IMRP can be applied to GWAS summary statistics of an exposure and an outcome obtained with overlapping or unique samples. To ensure the causal estimate is robust, we also applied a substantially different MR approach MRmix ^25^, an estimating equation approach that assumes 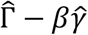 follows a normal mixture model. MRmix usually shows a good trade-off between bias and variance even with more than 50% invalid IVs ^25^. MRmix requires standardized summary statistics and IMRP does not. For a continuous trait, the effect size is rescaled by 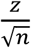, where z and n correspond to the z-score for an IV and the sample size, respectively, with its standard error 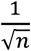. For a binary trait, the effect size is rescaled by 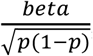, where beta and p correspond to the effect size and minor allele frequency of an IV, respectively. This standardizing procedure has been used in MRmix^25^.

**Figure 1.**
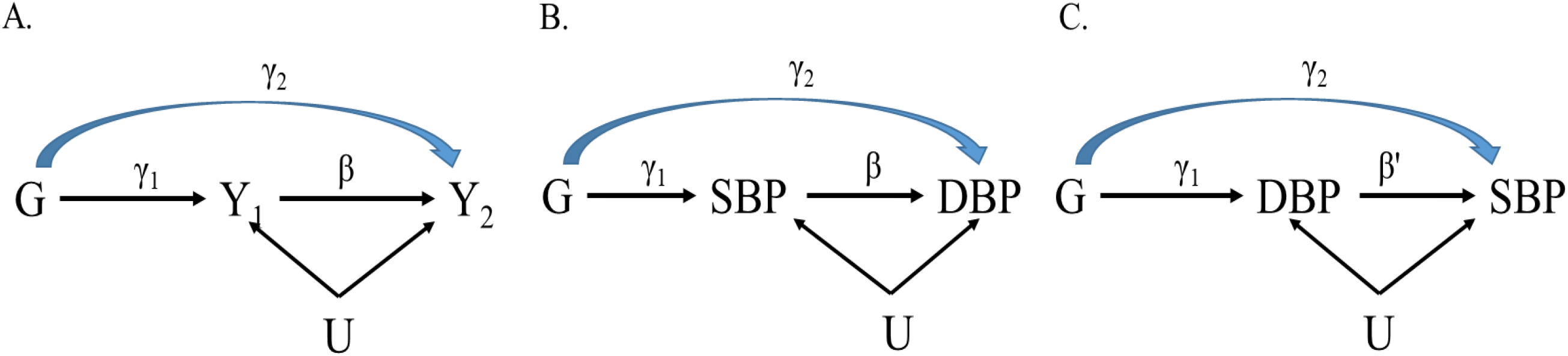
Path diagram in MR and Pleiotropic analysis. A. General path diagram in MR analysis. B and C. Path diagrams when SBP causally contributes to DBP and DBP causally contributes to SBP, respectively.

### GWAS of pleiotropy analysis for SBP and DBP

After performing a bi-directional MR analysis of SBP and DBP and estimating the causal effects of SBP on DBP and DBP on SBP, we extended the pleiotropy test *T*_*Pleio*_ to all 7.1M SNPs by fixing the causal effects estimated from IMRP analysis in the two causal paths, using UKB-ICBP summary statistics (**Figure 1B** and **C**). This is equivalent to performing GWAS for two new traits: BP_pleio1_ = DBP − *β* × SBP and BP_pleio2_ = SBP − *β*′ × DBP, where *β* and *β*′ are the estimated causal effects of SBP on DBP and DBP on SBP, respectively:

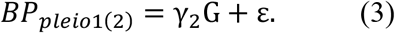

Equivalently, *T*_*Pleio*_ tests the null hypothesis *γ*_2_ = 0. Unlike MR analysis where the IVs are selected to be associated with exposure, we required the pleiotropy test *T*_*Pleio*_ for both BP_pleio1_ and BP_pleio2_ to be significant to declare a pleiotropic variant. We performed the same analysis for replication using MVP summary statistics. Meta-analysis of UKB-ICBP and MVP was further performed to increase statistical power and to identify additional variants.

We applied the LD score regression method^29^ to test for genomic inflation in the GWAS pleiotropy analysis. It is expected that BP_pleio1(2)_ will have a large genomic control inflation coefficient because of large sample sizes, genetic variants in high LD and a large number of BP variants^6^. We examined the degree of inflation from the intercept of the LD score regression.

### Novel locus definition

Novel loci were defined as genome wide significant pleiotropy variants > 1Mb away from known BP variants as well as LD r^2^ <0.1 with any known BP variants. Novel signals at a known locus were genome wide significant pleiotropy variants within 1 Mb of known BP variants as well as not being in LD with any known BP variants (*r*^2^ < 0.1) at the locus. The 1000G European ancestry data was used as the reference genetic data for LD calculation.

### Functional annotations

We evaluated all sentinel SNPs at novel loci for evidence of mediation of expression quantitative trait loci (eQTL) and splicing quantitative trait loci (sQTL) in all 44 tissues using the Genotype-Tissue Expression (GTEx) database. Following the method in Evangelou et al.^6^, a locus is annotated with a given eGene(sGene) only if the most significant eQTL(sQTL) SNP for the given eGene(sGene) is in high LD (*r*^2^ ≥ 0.8) with the sentinel SNP. We performed overall enrichment tests using the mediation and pleiotropic variants separately. We used DEPICT^30^ (Data-driven Expression Prioritized Integration for Complex Traits) to identify tissues and cells that are highly expressed at genes within the BP mediation and pleiotropic loci. We also used DEPICT to test for enrichment in gene sets associated with GO ontologyies, mouse knockout phenotypes and protein-protein interaction networks. We reported significant enrichments with a false discovery rate of 0.05. Analysis was done using the platform Complex-Traits Genetics Virtual Lab^31^.

### Genetic risk score (GRS) and pleiotropic Genetic Risk Scores (pGRS)

We constructed a traditional genetic risk score using independent genome wide significant BP variants from UKB-ICBP. We first constructed SBP and DBP weighted GRSs and then derived a single BP core GRS (cGRS) as the average of SBP and DBP GRSs. This approach has been previously used^6^ to estimate the combined effect of BP variants on BP, hypertension and CVD. Analogously, we constructed a pleiotropic genetic risk score (pGRS) using the variants detected in pleiotropy analysis. We first constructed BP_pleio1_ and BP_pleio2_ weighted GRSs and next derived the pGRS as the difference of BP_pleio1_ and BP_pleio2_ GRSs. Because most of the genetic variants associated with BP_pleio1_ and BP_pleio2_ demonstrated pleiotropy evidence, we termed this the pleiotropy genetic risk score. Noted that some SNPs contribute to both cGRS and pGRS because these variants are significantly associated with SBP or DBP, as well as BP_pleio1_ and BP_pleio1_. However, their weights represent their corresponding contributions to BP through the mediation and pleiotropy pathways. We performed linear regression analyses by jointly modeling cGRS and pGRS with and without adding the interaction of age*cGRS and age*pGRS on BP in the UKB data. We included the covariates of sex, age, BMI, geographical region and 10 genetic principal components in the linear regression analysis. The heritability explained by the cGRS and pGRS was calculated by the adjusted R^2^ in the linear regression adjusting out the covariates. Similarly, we performed logistic regression of cGRS, pGRS with and without age*cGRS and age*pGRS interactions on hypertension and cardiovascular events at baseline in the UKB data. The same covariates were included. We calculated the Nagelkerke’s R^2^ to quantify the goodness-of-fit of the prediction by the cGRS and pGRS^32^. We examined whether pGRS is able to predict additional variations of BP, hypertension and CVD after accounting for cGRS. We also examined the age-varying effects of cGRS and pGRS by testing interaction effects. Our analysis included 386,752 unrelated individuals of European ancestry with phenotypes measured at baseline. For comparison, we further constructed the PP weighted GRS (ppGRS) and performed the above analysis.

We assessed association of cGRS, pGRS and their interactions with age on BP in unrelated Africans (*n* = 7,904) and South Asians (*n* = 8,509) from the UKB to see whether BP– associated SNPs identified from GWAS predominantly in Europeans are also associated with BP in populations of non-European ancestry. Analysis was also performed for ppGRS. All analyses were performed using residuals after adjusting for sex, age, BMI, geographical region and 10 genetic principal components.

### Cross-trait lookups of novel loci

We supplied the index SNPs at the novel loci observed in UK Biobank-ICBP pleiotropic analyses to FUMA^33^ and GWAS catalog^34^ to investigate pleiotropy with non-BP traits, extracting all associations with P < 5 × 10^−8^ for all SNPs in high LD (r^2^ ≥0.8).

## Results

We present a bi-directional MR analysis of SBP and DBP using 1,125 and 1,183 independent genome-wide significant variants for SBP and DBP (P< 5×10^−8^) as genetic IVs obtained from the UKB-ICBP GWAS^6^. We standardized SBP and DBP and obtained an identical causal effect of SBP on DBP and DBP on SBP (0.864 ± 0.005 and 0.862 ± 0.005 by IMRP, respectively, **Supplementary Table 1**), which is significantly larger than the observed trait correlation 0.738 between SBP and DBP in UKB European subjects, with an estimated 74.8% of variation is the shared causal contribution between SBP and DBP. The causal estimates by MRmix were concordant (0.89 ± 0.012 and 0.90 ± 0.01, respectively, **Supplementary Table 1**). Among the genetic IVs, 43% of the variants had pleiotropic effects on SBP and DBP.

We next extended the pleiotropic effect analysis to search for variants by performing two GWAS of BP_pleio1_ and BP_pleio2_; the Manhattan and QQ plots are presented in **Figure 2**. The GC lambda value was 1.533 and LDSR intercept was 1.057 (0.013), with inflation ratio 4.23%, suggesting little inflation in the BP_pleio1_; a similar result was observed for BP_pleio2_. LD score regression analysis^29^ estimated 8.7% of the heritability arising from pleiotropic variants for BP_pleio1_. We calculated genetic correlations of BP_pleio1_ and BP_pleio2_ with SBP, DBP and PP using summary statistics (**Supplementary Table 2)**. BP_pleio1_ and BP_pleio2_ are genetically highly correlated (r_g_ =-0.902±0.006), also highly correlated with PP (r_g_ =-0.618±0.016 and 0.897±0.004) but less so with SBP or DBP. BP_pleio1_ is negatively correlated with SBP as is BP_pleio2_ with DBP; in contrast, PP is positively correlated with both SBP and DBP.

**Figure 2.**
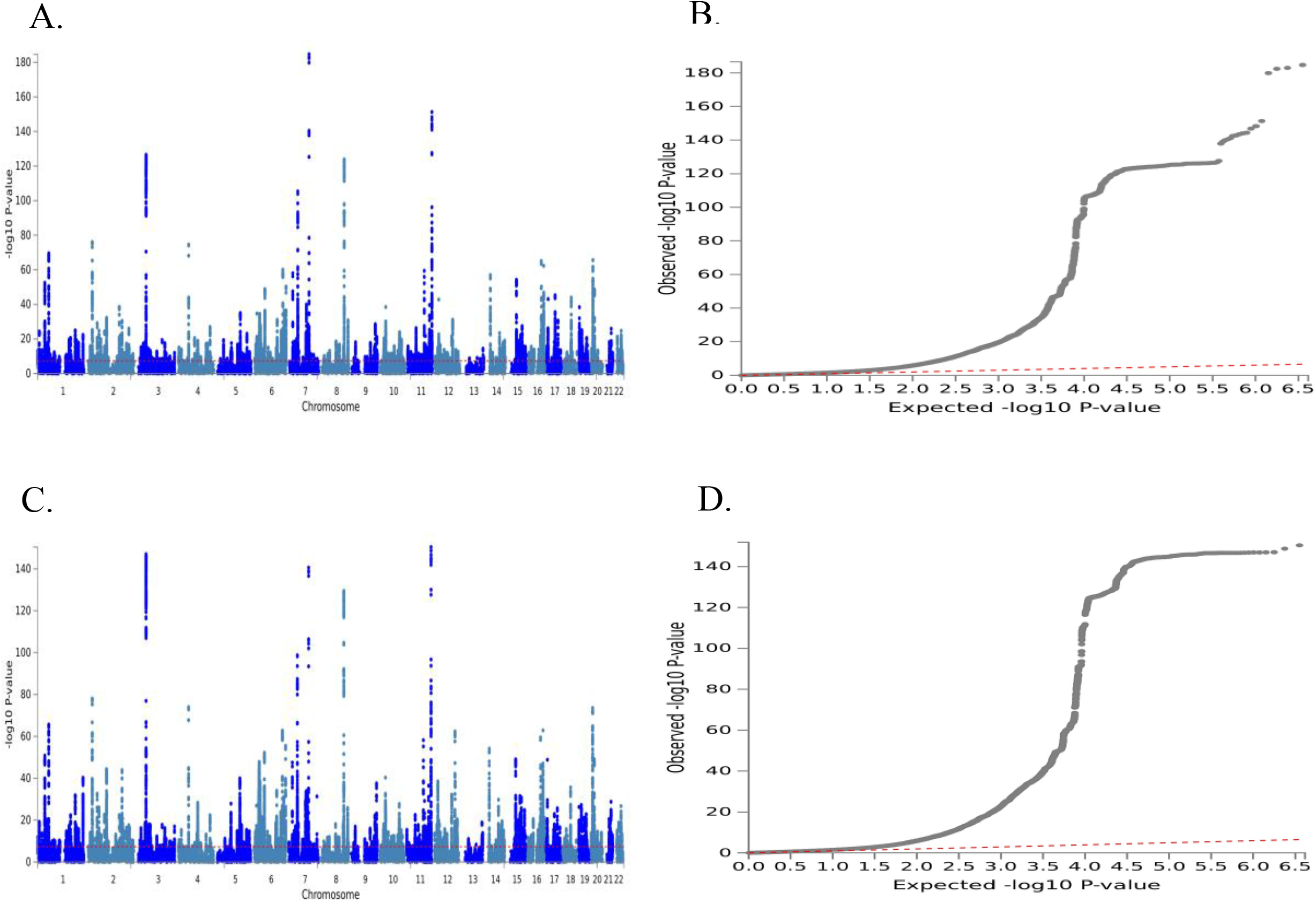
Manhattan and QQ plots for genome wide pleiotropy tests between SBP and DBP using UK Biobank-ICBP summary statistics. The GWAS of pleiotropy tests is equivalent to performing GWAS for two new traits: BP_pleio1_=DBP − *β* ∗ SBP and BP_pleio2_=SBP − *β*′ ∗ DBP, where *β* and *β*′ are the estimated causal effects of SBP on DBP and DBP on SBP, respectively. **A. and B**. Manhattan and QQ plots for BP_pleio2_. **C. and D**. Manhattan and QQ plots for BP_pleio1_. The horizontal line in Manhattans represents P-value=5×10^−8^. The top and bottom Manhattan plots are highly similar, indicating the consistence of the two directional MR analysis.

We observed 906 independent variants (r^2^ <0.1) reaching genome-wide significance in either BP_pleio1_ or BP_pleio2_ (P<5×10^−8^). To declare a variant as pleiotropic, we required one pleiotropy test P-value ≤5×10^−8^ and the other test P-value ≤0.05/906 by adjusting for multiple comparisons. We observed 815 independent pleiotropy variants with 91 variants genome wide significant also for either SBP or DBP. Among them, 234 (or 29%) were not detected by the univariate GWAS analysis of SBP, DBP or PP in the original UK Biobank+ICBP consortium^6^ (**Supplementary Table 3**). Among the 234 variants, 201 (or 86%) variants were not reported in any previous BP GWAS. In the set of associations, 163 variants in 124 loci were within a 1Mb region of previously reported known BP loci but were not in LD with known BP variants (r^2^ <0.1); the remaining 38 variants were at least 1Mb away from the previous reported known BP loci and resided at 35 loci; the corresponding locus zoom plots are presented in **Supplementary Figure 1**. We evaluated the associations of our sentinel SNPs at the 35 novel loci with other traits and disease using the GWAS Catalog^34^ and FUMA^33^. The GWAS Catalog and FUMA search of published GWAS showed that 29 of the 35 novel loci are also significantly associated with other traits, including lipid levels, cardiovascular-related outcomes, anthropometric traits, sleep traits, educational attainment, smoking, blood protein level and Schizophrenia (**Supplementary Table 4**).

We defined the variants with P-value of SBP < 5×10^−8^ and P-value of BP_pleio1_ >0.05/906, or P-value of DBP < 5×10^−8^ and P-value of BP_pleio2_ >0.05/906 into mediation variants, which resulted in 1,415 independent variants. We compared the SBP and DBP effect sizes for these mediation variants and observed that the mediation variants have the same effect directions for SBP and DBP (**Figure. 3**). In comparison, 71% of pleiotropic variants show opposite effect directions for SBP and DBP, indicating new discoveries.

**Figure 3.**
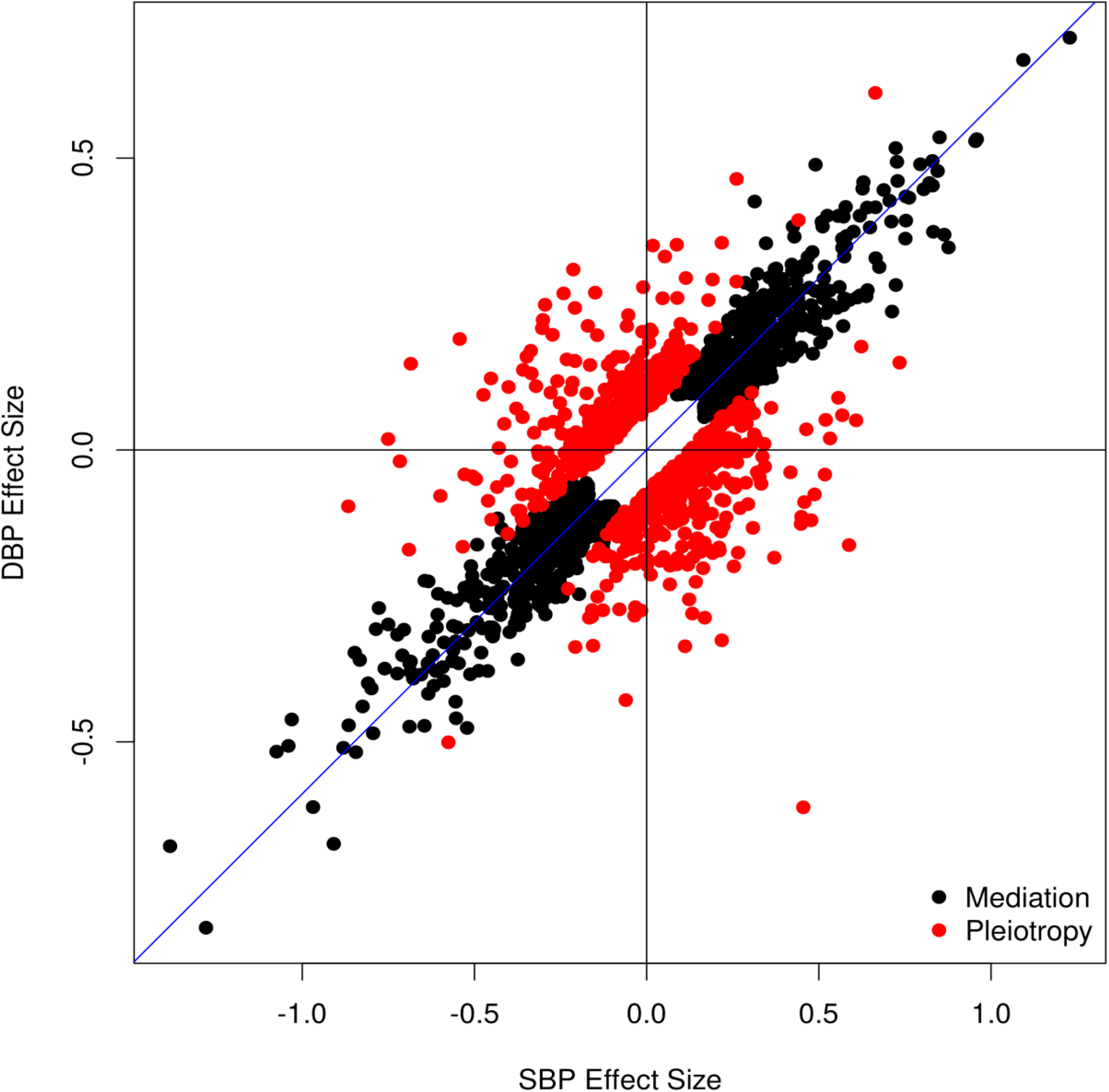
Comparison between the SBP and DBP effect sizes for mediation (black dots) and pleiotropic (red dots) variants in UKB-ICBP.

### Replication of novel signals in MVP

Since the BP summary statistics in MVP were obtained from multiethnic populations of non-Hispanic whites, non-Hispanic blacks, Hispanics, non-Hispanic Asians and non-Hispanic Native Americans, we performed MR analysis to estimate causal effect sizes between SBP and DBP in MVP, instead of using the causal effects estimated from UKB-ICBP. With a fewer number of IVs in MVP, we obtained relatively smaller but similar causal effect sizes (0.692 and 0.724) between SBP and DBP as compared to UKB-ICBP results, possibly due to the multiethnic samples in MVP (**Supplementary Table 1**). We then performed the pleiotropy test *T*_*Pleio*_ among the 201 novel variants (7 were not available in MVP). We examined how many novel variants significantly associated with BP_pleio1(2)_ (P<5×10^−8^) in UKB-ICBP also showed replication with corresponding BP_pleio1(2)_ at significance level P<0.05/201. We were able to replicate 23 variants (**Supplementary Table 3**), including 2 novel loci (rs12470661 and rs73937040, **Table 1**). When we released the replication significance level criterion at P=0.05, we were able to replicate 118 variants including 19 variants at 18 novel loci (**Table 1**), with 84 variants having both BP_pleio1_ and BP_pleio2_ P-values < 0.05 (**Supplementary Table 3**) and all the 118 variants having the same effect direction between UKB-ICBP and MVP: thus, our identified novel pleiotropic signals were replicable.

**Table 1.** The 17 novel BP loci identified by pleiotropic analysis.

| SNP | CHR | BP | UKB-ICBP<br>P_SBP | UKB-ICBP<br>P_DBP | UKB-ICBP<br>P_PP | UKB-ICBP<br>P<sub>pleio</sub><sup>a</sup> | MVP<br>P<sub>pleio</sub><sup>a</sup> | UKB-ICBP<br>-MVP P<sub>pleio</sub><sup>a</sup> | Genes |
| --- | --- | --- | --- | --- | --- | --- | --- | --- | --- |
| rs11162906 | 1 | 80500074 | $3.19 \times 10^{-2}$ | $1.75 \times 10^{-2}$ | $1.78 \times 10^{-6}$ | $2.41 \times 10^{-9}$ | $1.53 \times 10^{-3}$ | $3.54 \times 10^{-11}$ | *AC098657.2* |
| rs17713879 | 2 | 254215 | $5.19 \times 10^{-2}$ | $2.66 \times 10^{-2}$ | $2.05 \times 10^{-5}$ | $3.75 \times 10^{-8}$ | $8.92 \times 10^{-4}$ | $1.35 \times 10^{-10}$ | *SH3YL1* |
| rs3136302 | 2 | 48021379 | $4.08 \times 10^{-1}$ | $5.84 \times 10^{-5}$ | $9.62 \times 10^{-6}$ | $2.96 \times 10^{-11}$ | $4.48 \times 10^{-2}$ | $8.30 \times 10^{-12}$ | *MSH6* |
| rs6735304 | 2 | 101617631 | $4.18 \times 10^{-2}$ | $2.38 \times 10^{-2}$ | $1.43 \times 10^{-6}$ | $1.47 \times 10^{-8}$ | $6.21 \times 10^{-4}$ | $6.36 \times 10^{-11}$ | *RPL31/TBC1D8* |
| rs12470661 | 2 | 232060050 | $5.09 \times 10^{-2}$ | $3.07 \times 10^{-3}$ | $6.68 \times 10^{-7}$ | $5.33 \times 10^{-11}$ | $1.97 \times 10^{-4}$ | $4.55 \times 10^{-14}$ | *HTR2B/ARMC9* |
| rs10947978 | 6 | 41471608 | $7.01 \times 10^{-1}$ | $9.89 \times 10^{-6}$ | $1.44 \times 10^{-5}$ | $2.65 \times 10^{-11}$ | $1.84 \times 10^{-2}$ | $3.26 \times 10^{-12}$ | *LINC01276* |
| rs56098119 | 6 | 90296727 | $7.63 \times 10^{-2}$ | $1.38 \times 10^{-2}$ | $6.81 \times 10^{-6}$ | $1.69 \times 10^{-8}$ | $2.89 \times 10^{-2}$ | $1.77 \times 10^{-9}$ | *ANKRD6* |
| rs150953973 | 6 | 120780033 | $2.98 \times 10^{-1}$ | $9.7 \times 10^{-4}$ | $4.01 \times 10^{-6}$ | $3.54 \times 10^{-9}$ | $1.64 \times 10^{-2}$ | $1.85 \times 10^{-10}$ | *RNU6-214P* |
| rs180271 | 7 | 93539479 | $6.11 \times 10^{-1}$ | $1.72 \times 10^{-4}$ | $2.63 \times 10^{-5}$ | $3.69 \times 10^{-9}$ | $2.05 \times 10^{-3}$ | $2.92 \times 10^{-11}$ | *GNGT1* |
| rs11989271 | 8 | 122632611 | $1.51 \times 10^{-1}$ | $8.47 \times 10^{-4}$ | $7.80 \times 10^{-7}$ | $1.14 \times 10^{-10}$ | $9.37 \times 10^{-3}$ | $1.38 \times 10^{-11}$ | *HAS2* |
| rs10868842 | 9 | 73119085 | $1.57 \times 10^{-1}$ | $3.58 \times 10^{-4}$ | $9.71 \times 10^{-6}$ | $1.37 \times 10^{-11}$ | $4.07 \times 10^{-3}$ | $2.54 \times 10^{-13}$ | *LINC00583* |
| rs12768143 | 10 | 22808844 | $3.92 \times 10^{-2}$ | $5.03 \times 10^{-3}$ | $8.61 \times 10^{-8}$ | $9.18 \times 10^{-11}$ | $1.58 \times 10^{-2}$ | $2.39 \times 10^{-11}$ | *PIP4K2A* |
| rs1343676 | 12 | 33537387 | $7.19 \times 10^{-1}$ | $2.03 \times 10^{-7}$ | $8.65 \times 10^{-7}$ | $9.56 \times 10^{-15}$ | $4.04 \times 10^{-4}$ | $3.04 \times 10^{-16}$ | *SYT10* |
| rs7322054 | 13 | 38246708 | $6.71 \times 10^{-1}$ | $1.99 \times 10^{-7}$ | $7.76 \times 10^{-4}$ | $1.04 \times 10^{-11}$ | $1.01 \times 10^{-2}$ | $5.86 \times 10^{-13}$ | *TRPC4* |
| rs61972411 | 13 | 100602630 | $2.82 \times 10^{-4}$ | $6.59 \times 10^{-1}$ | $1.87 \times 10^{-7}$ | $1.45 \times 10^{-8}$ | $1.89 \times 10^{-2}$ | $2.23 \times 10^{-9}$ | *LOC101927437* |
| rs62621400 | 15 | 101718239 | $2.25 \times 10^{-1}$ | $1.70 \times 10^{-3}$ | $1.94 \times 10^{-5}$ | $3.76 \times 10^{-9}$ | $1.64 \times 10^{-3}$ | $2.34 \times 10^{-11}$ | *CHSY1* |
| rs116643984 | 15 | 101791212 | $1.64 \times 10^{-1}$ | $7.73 \times 10^{-5}$ | $5.14 \times 10^{-8}$ | $4.04 \times 10^{-13}$ | $6.86 \times 10^{-3}$ | $1.60 \times 10^{-14}$ | *CHSY1* |
| rs73937040 | 18 | 3258733 | $7.08 \times 10^{-2}$ | $2.07 \times 10^{-2}$ | $7.77 \times 10^{-6}$ | $4.67 \times 10^{-8}$ | $2.06 \times 10^{-6}$ | $1.17 \times 10^{-12}$ | *MYL12A/MYL12B* |
| rs146827176 | 20 | 35169916 | $6.48 \times 10^{-1}$ | $3.10 \times 10^{-6}$ | $1.01 \times 10^{-3}$ | $2.13 \times 10^{-9}$ | $5.08 \times 10^{-3}$ | $3.82 \times 10^{-11}$ | *LOC101926987/MYL9* |

<sup>a</sup>The p-values of pleiotropy test for BP<sub>pleio1</sub> and BP<sub>pleio2</sub> were consistent in general and we reported the lesser one here. The detailed summary statistics and corresponding P values were listed in Supplementary **Table 3**.

We further examined the BP_pleio1(2)_ effect size consistency of the 815 independent pleiotropic variants between UKB-ICBP and MVP. For comparison, we also examined the SBP and DBP effect size consistency of the independent 1,415 mediation variants between UKB-ICBP and MVP. We observed that the BP_pleio1(2)_ effect sizes have higher correlations for the pleiotropic variants than the mediation variants (**Figure. 4**, correlation 0.90 vs 0.74), even after exclusion of 2 significant outliers (rs113081691 and rs17057329). We observed that 96% of pleiotropic variants have consistent effect directions between UKB-ICBP and MVP. We then performed inverse variance weighted meta-analysis to combine the summary statistics of BP_pleio1(2)_ for UKB-ICBP and MVP. The combined pleiotropy evidence was further strengthened for the 118 novel variants (**Table 1** and **Supplementary 3)**.

**Figure 4.**
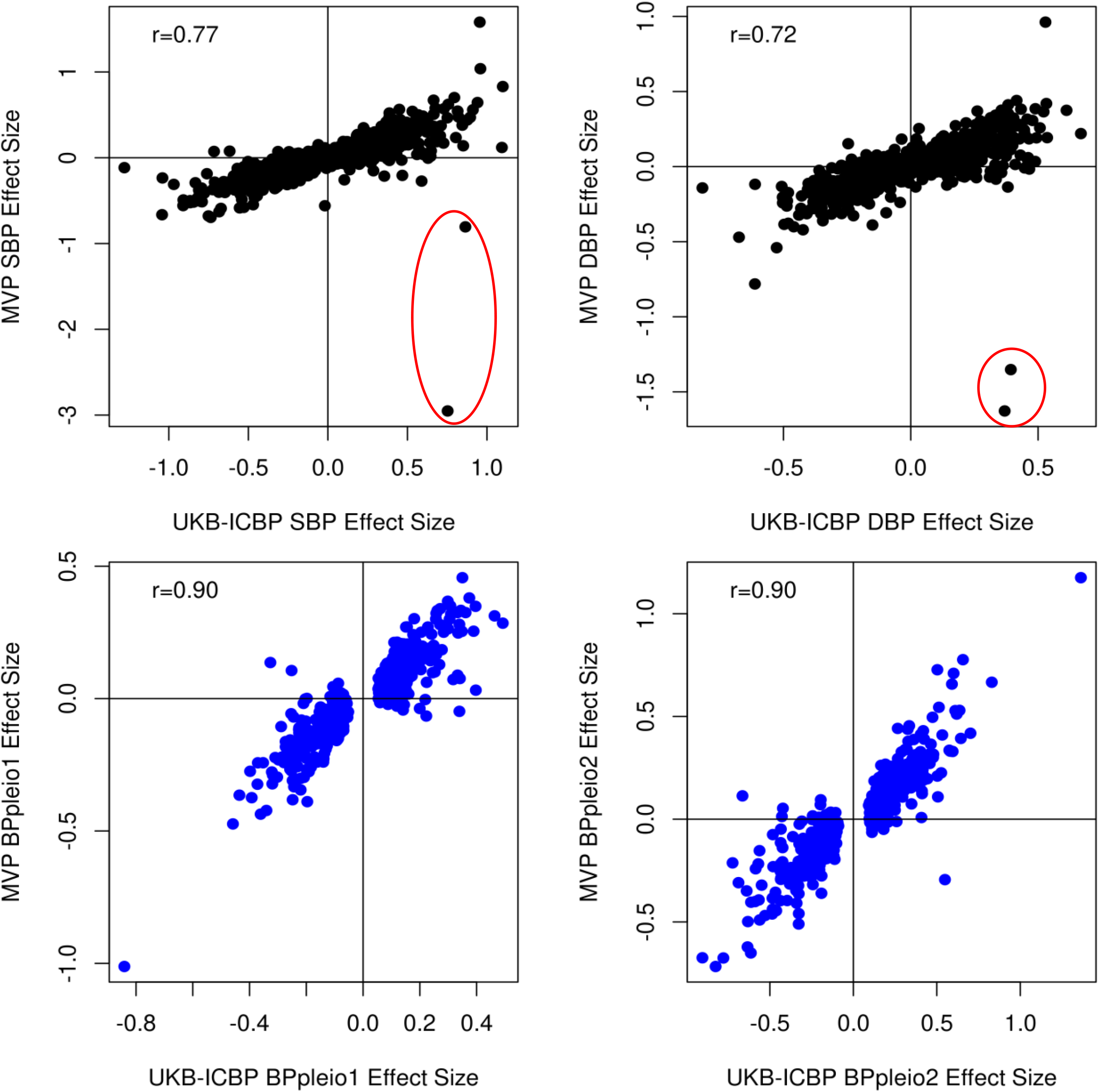
Comparison between the effect sizes between UKB-ICBP and MVP. **A**. SBP effect sizes of mediation variants. **B**. DBP effect sizes of mediation variants. **C**. BP_pleio1_ effect sizes for pleiotropic variants. **D**. BP_pleio2_ effect sizes for pleiotropic variants. The two variants rs113081691 and rs17057329 in the red circles in **A**. and **B**. represent substantial different effect sizes of SBP and DBP between UKB-ICBP and MVP, likely driven by multi-ethnic samples from MVP.

### Functional annotations

We performed expression quantitative trait locus (eQTL) analysis using GTEx data. Among the 35 novel loci listed in **Supplementary Table 3**, we identified 26 with expression quantitative trait locus (eQTL) (**Supplementary Table 5**) and 11 with Splicing Quantitative Trait Loci (sQTLs) (**Supplementary Table 6**). The eQTLs were most often enriched in arterial tissues, followed by adipose, heart and nerve tibial tissues. SNP rs17713879 is an eQTL affecting expression of the *SH3YL1* and *ACP1* genes in 34 tissues and is also a sQTL affecting splicing of these two genes in 50 tissues. SNP rs112500920 is an eQTL affecting expression of several genes, including *EFL1* and *AB3B2*, in multiple tissues, notably adipose and arterial tissues. SNP rs12478520 is an eQTL affecting expression of multiple genes, including *C2orf72, HTR2B, ARMC9* and *PSMD1*.

In the UKB-ICBP data, we identified 815 independent pleiotropic variants. We also observed 1,451 independent mediation variants,. We assessed tissue enrichment of BP loci using DEPICT^30^ at a false discovery rate (FDR) < 5% but separated 1,415 mediation from 815 pleiotropic variants in the analysis. DEPICT identified enrichment across 43 and 51 tissues and cells using mediation and pleiotropic variants, respectively (**Supplementary Table 7**). The enriched tissues are highly similar (correlation=0.78) but there are also notable differences (**Supplementary Figure 2a**). Enrichment was greatest for arteries in the cardiovascular system for both mediation and pleiotropic variants (P= 1.28 × 10^−3^ and 2.19 × 10^−11^, respectively). In general, enrichment observed for mediation variants were also observed for pleiotropic variants, but not vice versa. For example, heart related tissues, aortic valves and atrial appendage were enriched for pleiotropic variants (P< 1.38 × 10^−4^) but not for mediation variants (**Supplementary Table 7**). Pathway enrichments for mediation variants and pleiotropic variants were less well correlated (correlation=0.51, **Supplementary Figure 2b**). Pleiotropic variants were enriched in many molecular pathways that were missed by mediation variants, including response to hypoxia, oxygen levels, basement membrane, and renal system development (P<8.66×10^−7^, **Supplementary Table 8**). In contrast, negative regulation of transcription from RNA polymerase II promoter, histone deacetylase binding, hormone receptor binding and Ras protein signal transduction, among others, were only enriched by mediation variants (P<6.09×10^−7^, **Supplementary Table 8**).

Evaluation of enriched mouse knockout phenotype terms by both mediation and pleiotropic variants implicated abnormal cardiovascular physiology, disorganized myocardium, abnormal vascular branching morphogenesis and organogenesis, among others. However, pleiotropic variant-enriched mouse phenotypes include abnormal kidney morphology, impaired wound healing, dilated heart right ventricle, abnormal aorta morphology, increased systemic arterial SBP and increased body weight (P< 1.65 × 10^−7^). Mediation variants-enriched mouse phenotypes include pericardial edema, wavy neural tube, and decreased systemic arterial BP (P<1.83×10^−4^, **Supplementary Table 8**). Common protein–protein interaction subnetwork enrichments for both mediation and pleiotropic variants include LAMA1, ITGB1, WNT1 and many SMAD subnetworks. Pleiotropic variant enriched top significant subnetworks include the HSPG2, TGM2, MMP9, FBN1 and DDR1 subnetworks (P<1.76× 10^−7^), as compared to mediation variant enriched subnetworks, SRC, YWHAQ and RAPGEF1 (**Supplementary Table 9**).

### Improved prediction of BP, hypertension and CVD by including pleiotropic variants

Polygenic scores derived from multiple related traits can improve prediction of outcomes ^35-38^. Chasman et al. decomposed a GRS into nearly independent components relative to biological mechanisms inferred from pleiotropic relationships ^39^ while Udler et al. factorized the genetic association matrix according to different classes of genetic variants relative to traits ^40^. However these approaches do not use the discovered pleiotropic variants directly. Here we construct a traditional BP GRS using all independent 1,615 BP variants from the UKB-ICBP, which we term the core genetic risk score (cGRS) (see **Material and Methods**). We further constructed a pGRS using the 906 variants associated with BP_pleio1_ or BP_pleio2_, which is a genetic risk score from pleiotropic variants. We jointly modeled cGRS and pGRS adjusting for age, gender, BMI and 10 principal components, and observed that pGRS significantly predicted BP traits, as well as risk of HTN and CVD, conditional on cGRS in all models (**Table 2A and Figure 5**) in the UKB European ancestry subjects. The cGRS captured 5.91%, 6.09% and 2.23% SBP, DBP and PP heritability excluding pGRS and 7.13%, 6.75% and 7.27% including pGRS, or a 1.11-to 3.26-fold increase. Similarly, Nagelkerke’s R^2^ for HTN and CVD was 4.71% and 0.47% excluding pGRS and 5.14% and 0.53% including pGRS, representing a 1.09- and 1.14-fold increase. We observed odds ratios (ORs) of 1.64 and 1.15 for individual cGRS and pGRS on the risk of HTN (*P* < 1 × 10^−300^), respectively. The observed ORs of individual cGRS and pGRS for CVD were 1.21 and 1.06 (*P* = 1.71 × 10^−231^ and *P* = 2.74 × 10^−25^), respectively, and increased to 1.66 and 1.19 (*P* = 9.83 × 10^−97^ and *P* = 1.28 × 10^−13^) when comparing the upper versus lower quantiles of the cGRS and pGRS, respectively. However, the odds ratios were further increased to 6.78 and 2.44 (*P* < 1 × 10^−300^ and *P* = 1.15 × 10^−39^) for HTN and CAD, respectively, when comparing the top decile and quintile with bottom decile and quintile of cGRS and pGRS (**Figure 5**). We observed a clear advantage of including pGRS over cGRS only (**Figure 5**). It has been suggested that there are gene and age interactions that contributing to blood pressure and hypertension^16; 17; 20^. However, the detected interactions are limited because of low statistical power. We thus examined the interaction effects between age and cGRS and pGRS. After including the interactions of age and cGRS and pGRS the main effects for cGRS and pGRS on BP, HTN and CVD were unchanged. However, we observed a significant interaction effect of age and cGRS for all BP traits and HTN but not CVD. The interaction of age and pGRS significantly contributed to all BP traits, HTN and CVD (P value between 3.0×10^−2^ and 7.74×10^−29^, **Table 2A)**.

**Table 2:**
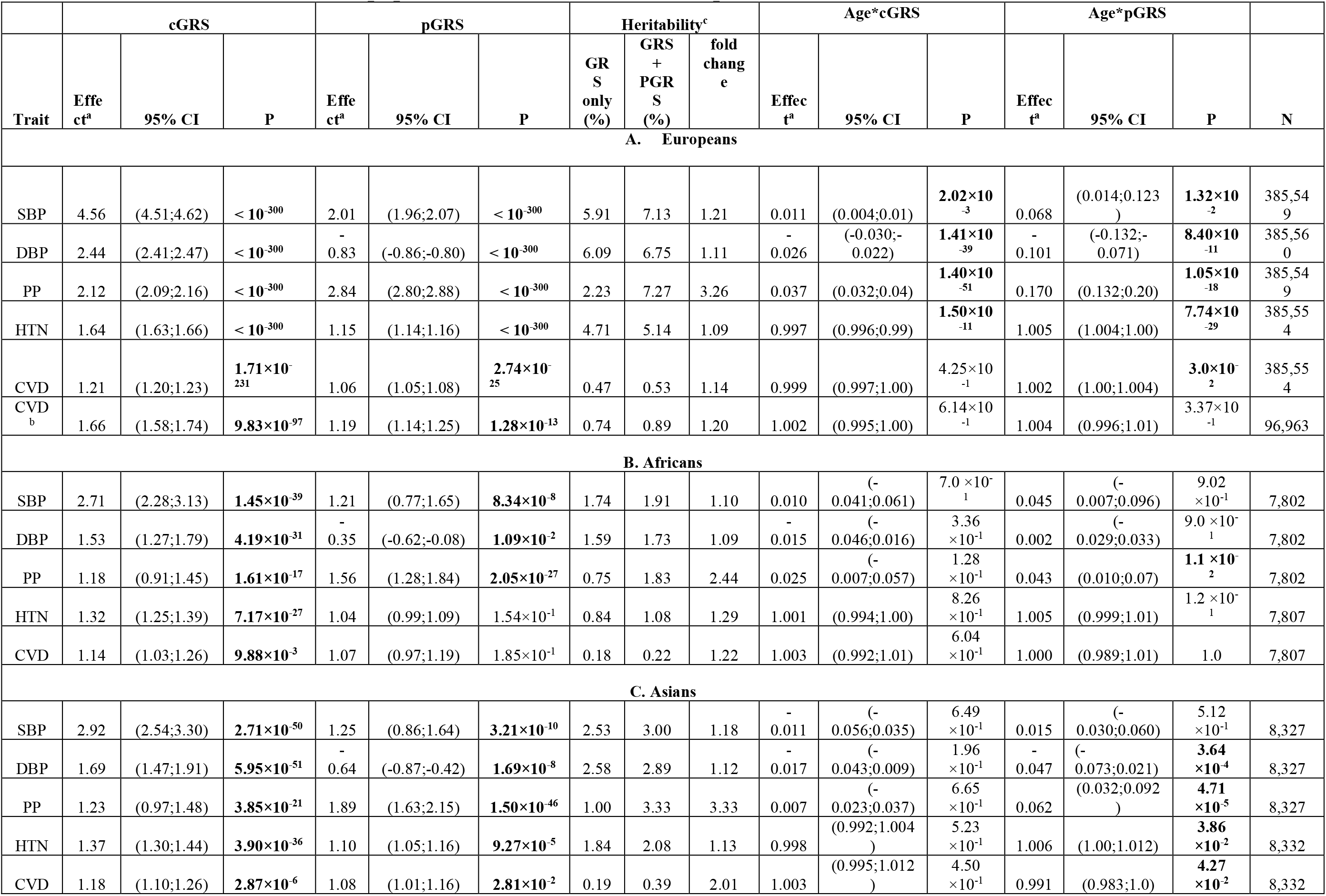
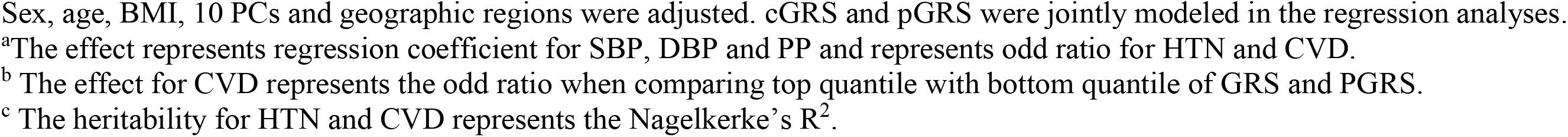
Associations of the cGRS, pGRS and their interactions with age on blood pressure traits, hypertension and cardiovascular events in unrelated populations in UK Biobank European, African and Asian descents.

**Figure 5.**
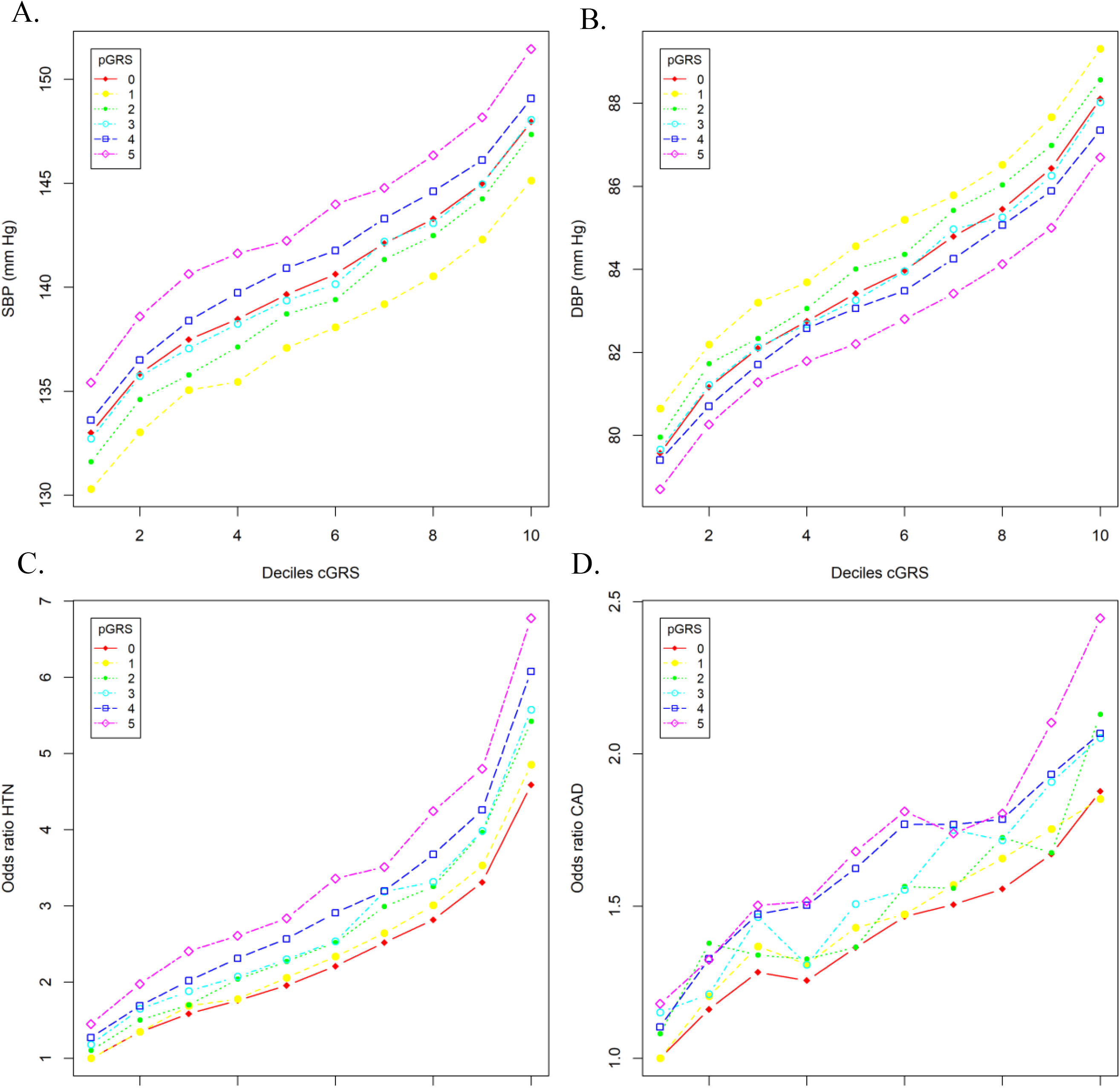
Relationship the core genetic risk score (cGRS) and pleiotropic genetic risk score (pGRS) with blood pressure, risk of hypertension and cardiovascular disease in UK Biobank. (A) sex adjusted mean systolic blood pressure (SBP); (B) sex adjusted mean diastolic blood pressure; (C) odds ratios of hypertension (HTN) and (D) odds ratios of cardiovascular disease (CVD). cGRS was calculated in every decile and pGRS was calculated every quintile. Odds ratios were calculated by comparing each of the cGRS deciles and pGRS quintiles with the lowest decile and twentieth. The curves with pGRS=0 represent the models without including pGRS.

### Extension to other ancestries

We examined associations with BP and CVD of the above defined European cGRS and pGRS in unrelated African (N=7,904) and South Asian (N=8,509) subjects in the UKB (**Table 2. B, C**). Although sample sizes were much smaller than among UKB European subjects, the cGRS is significantly associated with SBP, DBP, PP, HTN and CVD in both the UKB African- and Asian ancestry subjects. In the UKB African ancestry individuals, including the pGRS results in a 1.10-to 2.44-fold increase of SBP, DBP and PP heritability. Nagelkerke’s R^2^ for HTN and CVD has 1.23- and 1.22-fold increases, respectively. Similar increments in the UKB Asian cohort is also observed (**Table 2. C**). Significant interactions of age and pGRS were again observed for BP traits and CVD in UKB Asians (**Table 2. C**).

### Comparison between BP_pleio1_(BP_pleio2_) and PP

We noted PP is genetically positively correlated with both SBP and DBP but BP_pleio1_ or BP_pleio2_ is negatively correlated with SBP and DBP (**Supplementary Table 2**). Among the 815 pleiotropic variants, 274 were genome wide significant with either SBP or DBP and 467 with PP. We then calculated the GRS of PP (GRS_PP_) in UKB using the independent variants associated with PP by the effect sizes estimated in the UBK-ICBP data. We observed that the GRS_PP_ is positively correlated with both the GRSs of SBP and DBP. In comparison, the pGRS is again oppositely correlated with the GRSs of SBP and DBP (**Supplementary Table 2**). We next compared BP variances explained by the GRS_PP_ and pGRS in UKB. In general, GRS_PP_ and pGRS can account for similar amounts of BP, HTN and CVD variation (**Supplementary Table 9**). However, when we included cGRS, pGRS and GRS_PP_ in a regression model, we observed that all three genetic risk scores significantly predicted variation in BP, HTN and CVD in UKB European subjects (**Supplementary Table 9**), suggesting pGRS and GRS_PP_ identify different aspects of trait variations.

### Mendelian Randomization of BP on CAD, MI and stroke

We downloaded published GWAS summary statistics for CAD^41^, MI^42^ and stroke^43^ and performed MR analysis of SBP, DBP, PP, BP_pleio1_ and BP_pleio2_ on CAD, MI and Stroke (**Table** 3). For SBP and DBP, IVs were the genetic variants genome wide significantly associated with SBP and DBP but with no pleiotropic evidence. For BP_pleio1_ and BP_pleio2_, IVs were the genetic variants associated with BP_pleio1_ and BP_pleio2_. For PP, we selected all variants independently associated with PP as the IVs. The effect size and standard error of an IV for BP_pleio1_ or BP_pleio2_ were the corresponding numerator and denominator of the test statistic *T*_*Pleio*_ in equation (2). As expected, SBP, DBP and PP causally contributed to CAD, MI and stroke. The new trait - BP_pleio_ - also causally contributed to CAD, MI and stroke, suggesting a primary causal pleiotropy pathway that is not associated with BP mediation pathway directly contributing to the outcomes (**Table 3**). The estimated ORs ranged from 1.66 to 1.85 per SD unit increase in BP on the three outcomes using mediation variants and ranged from 1.13 to 1.48 using pleiotropic variants. Our analysis identified 1 to 22% IVs demonstrating pleiotropic effects for BP and the three clinical outcomes (**Table 3**).

**Table 3.**
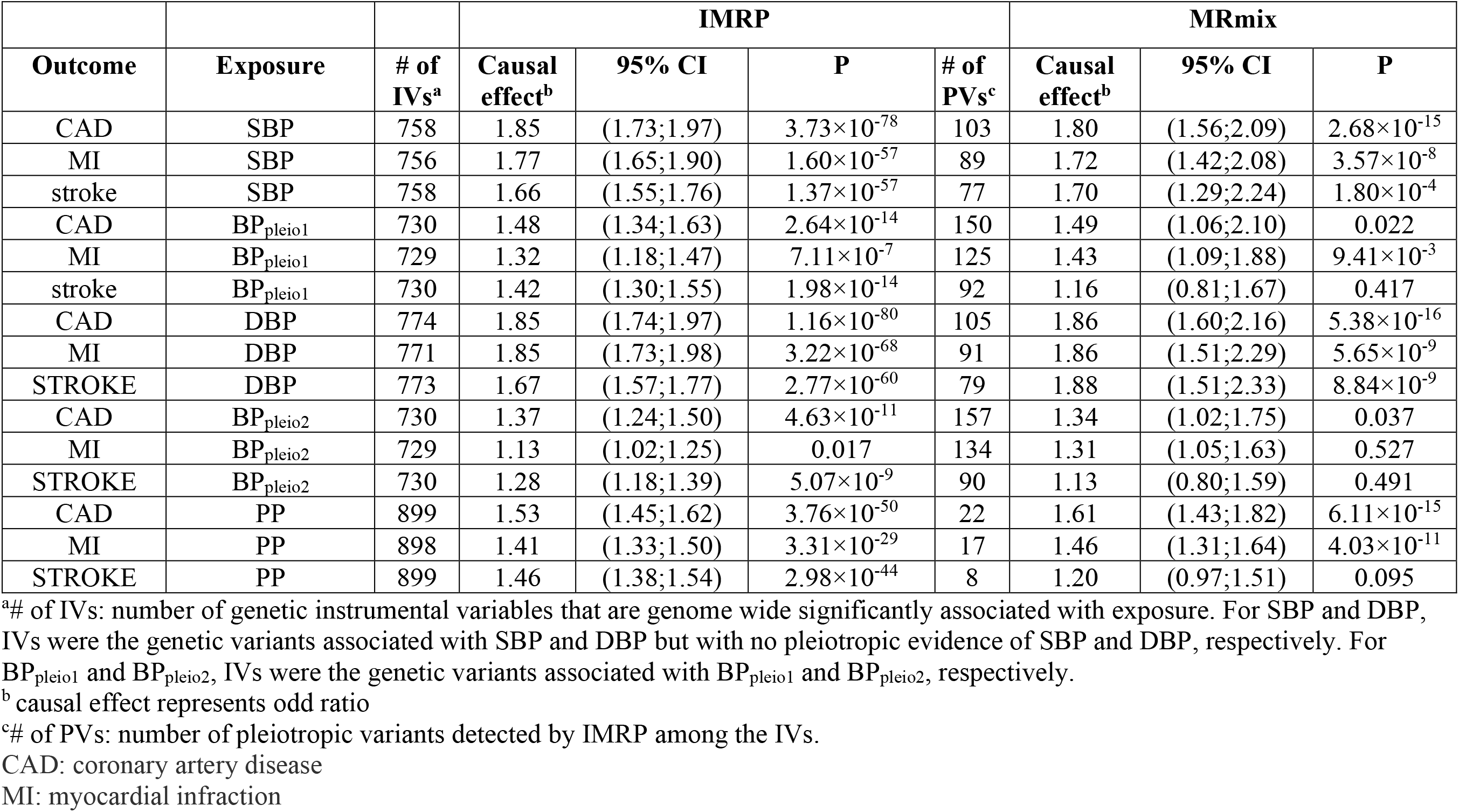
Using mediation variants and pleiotropy variants separately in MR analysis with outcomes: CAD, MI and STROKE.

| Outcome | Exposure | # of IVs<sup>a</sup> | IMRP |  |  |  | MRmix |  |  |
| --- | --- | --- | --- | --- | --- | --- | --- | --- | --- |
|  |  |  | Causal effect<sup>b</sup> | 95% CI | P | # of PVs<sup>c</sup> | Causal effect<sup>b</sup> | 95% CI | P |
| CAD | SBP | 758 | 1.85 | (1.73;1.97) | $3.73 \times 10^{-78}$ | 103 | 1.80 | (1.56;2.09) | $2.68 \times 10^{-15}$ |
| MI | SBP | 756 | 1.77 | (1.65;1.90) | $1.60 \times 10^{-57}$ | 89 | 1.72 | (1.42;2.08) | $3.57 \times 10^{-8}$ |
| stroke | SBP | 758 | 1.66 | (1.55;1.76) | $1.37 \times 10^{-57}$ | 77 | 1.70 | (1.29;2.24) | $1.80 \times 10^{-4}$ |
| CAD | BP<sub>pleio1</sub> | 730 | 1.48 | (1.34;1.63) | $2.64 \times 10^{-14}$ | 150 | 1.49 | (1.06;2.10) | 0.022 |
| MI | BP<sub>pleio1</sub> | 729 | 1.32 | (1.18;1.47) | $7.11 \times 10^{-7}$ | 125 | 1.43 | (1.09;1.88) | $9.41 \times 10^{-3}$ |
| stroke | BP<sub>pleio1</sub> | 730 | 1.42 | (1.30;1.55) | $1.98 \times 10^{-14}$ | 92 | 1.16 | (0.81;1.67) | 0.417 |
| CAD | DBP | 774 | 1.85 | (1.74;1.97) | $1.16 \times 10^{-80}$ | 105 | 1.86 | (1.60;2.16) | $5.38 \times 10^{-16}$ |
| MI | DBP | 771 | 1.85 | (1.73;1.98) | $3.22 \times 10^{-68}$ | 91 | 1.86 | (1.51;2.29) | $5.65 \times 10^{-9}$ |
| STROKE | DBP | 773 | 1.67 | (1.57;1.77) | $2.77 \times 10^{-60}$ | 79 | 1.88 | (1.51;2.33) | $8.84 \times 10^{-9}$ |
| CAD | BP<sub>pleio2</sub> | 730 | 1.37 | (1.24;1.50) | $4.63 \times 10^{-11}$ | 157 | 1.34 | (1.02;1.75) | 0.037 |
| MI | BP<sub>pleio2</sub> | 729 | 1.13 | (1.02;1.25) | 0.017 | 134 | 1.31 | (1.05;1.63) | 0.527 |
| STROKE | BP<sub>pleio2</sub> | 730 | 1.28 | (1.18;1.39) | $5.07 \times 10^{-9}$ | 90 | 1.13 | (0.80;1.59) | 0.491 |
| CAD | PP | 899 | 1.53 | (1.45;1.62) | $3.76 \times 10^{-50}$ | 22 | 1.61 | (1.43;1.82) | $6.11 \times 10^{-15}$ |
| MI | PP | 898 | 1.41 | (1.33;1.50) | $3.31 \times 10^{-29}$ | 17 | 1.46 | (1.31;1.64) | $4.03 \times 10^{-11}$ |
| STROKE | PP | 899 | 1.46 | (1.38;1.54) | $2.98 \times 10^{-44}$ | 8 | 1.20 | (0.97;1.51) | 0.095 |

<sup>a</sup># of IVs: number of genetic instrumental variables that are genome wide significantly associated with exposure. For SBP and DBP, IVs were the genetic variants associated with SBP and DBP but with no pleiotropic evidence of SBP and DBP, respectively. For BP<sub>pleio1</sub> and BP<sub>pleio2</sub>, IVs were the genetic variants associated with BP<sub>pleio1</sub> and BP<sub>pleio2</sub>, respectively.
<sup>b</sup> causal effect represents odd ratio
<sup>c</sup># of PVs: number of pleiotropic variants detected by IMRP among the IVs.
CAD: coronary artery disease
MI: myocardial infraction

### Meta-analysis of UKB-ICBP and MVP

We performed the inverse variance weighted meta-analysis of pleiotropy test to combine UKB-ICBP and MVP. We observed an additional 219 novel signals with either BP_pleio1_ or BP_pleio2_ at a significance level 5×10^−8^, including an additional 40 novel loci (**Supplementary Table 10**).

## Discussion

Our analysis of GWAS summary statistics from over 1 million subjects have here revealed important aspects of the genetic architecture of the two principle and highly correlated BP traits, SBP and DBP. The bi-directional MR analysis of SBP and DBP demonstrated that 1 SD unit increase of SBP leads to 0.86 SD unit increase of DBP, and vice versa, indicating that SBP and DBP share 74.8% of its variation. We assume these arise from common genetic factors and common biological mechanisms between SBP and DBP. This shared causal contribution is substantially higher than the 55.6% estimated by phenotype correlation analysis of SBP and DBP in UKB European ancestry subjects. We then identified the genetic variants that impact SBP and DBP through two different paths: 1) mediation path (either from SBP to DBP or vice versa) and departure from the mediation path (pleiotropic path, **Figure 1**). We defined the variants contributing to BP traits through the pleiotropic path as pleiotropic variants, which have different biological process from the BP variants through the mediation path. We observed that most of the BP variants identified through SBP or DBP univariate associations were mediation variants and would be expected to be discovered in either the SBP or DBP GWAS. By examining the variants departing from the mediation paths, we identified 815 independent variants demonstrating pleiotropy evidence in the original UK Biobank + ICBP consortium data^6^, of which 201 were undetected by univariate GWAS of SBP, DBP or PP in literature. Replication analysis in the MVP confirmed 118 of the 201 novel variants, including the 18 novel loci (**Table 1 and Supplementary Table 3)**. Pleiotropic variants often demonstrated an effect size opposite in direction for SBP and DBP and yet contributed 8.71% heritability of the newly defined BP trait (**Figure 3**). The effect sizes of pleiotropic variants also demonstrated a higher correlation than that of the mediation variants between UKB-ICBP and MVP, suggesting that the pleiotropic variants may be more transferable across ethnic populations. BP_pleio1_ and BP_pleio2_ were highly correlated with PP (ρ ≥0.62) in UKB whites, taken as an indicator of arterial stiffness and considered as an independent risk factor for CVD^44^. However, BP_pleio1_ and BP_pleio2_ were less correlated than PP with either SBP or DBP, which is consistent with pGRS being less correlated than GRS_PP_ with either SBP or DBP-defined GRS (**Supplementary Table 2**). Thus, BP_pleio1_ or BP_pleio2_ represent a different risk factor of CVD from PP. This is consistent with the finding that GRSs of SBP, DBP, PP and BP_pleio_ all contribute to risk of CAD, Stroke and MI in the MR analysis (**Table 3**). Thus, our results clearly suggest that a substantial fraction of BP variants affect both SBP and DBP through pleiotropic effects. By combining UKB-ICBP and MVP, we identified an additional novel 219 variants with pleiotropic evidence, including 40 novel loci, although independent replication of these latter results are warranted (**Supplementary Table 10**). Thus, pleiotropic variant searches in existing datasets can identify many new BP genes.

In addition to the traditional BP GRS^3; 6^, which we termed the core genetic risk score cGRS, the pleiotropic genetic risk score pGRS independently predicted BP, HTN and CVD outcomes (**Table 2**) in the UK Biobank European ancestry subjects. Additionally, including the pGRS led to substantial increments in heritability explained for BP traits (**Table 2 and Figure 5**). Although we observed consistent opposite directional effects of pGRS for SBP and DBP in UK Biobank European-, African- and Asian-ancestry subjects, the prediction of HTN and CVD risk was significantly improved by including pGRS (**Figure 5 C and D**). The cGRS and pGRS defined in European participants both consistently and significantly predicted BP, HTN and CVD in UK Biobank Africans and Asians, suggesting that pGRS is able to improve prediction accuracy across populations. Recent studies have suggested that cGRS models alone have modest improvement of predictive accuracy for CAD^45-47^. The principle outcome of this set of analyses, therefore, demonstrates that adding pGRS significantly improves the prediction model over cGRS alone. This approach of constructing polygenic risk scores is conceptually different from existing approaches using multiple related traits^35-38^ and can be generalized to other diseases by incorporating multiple disease-related traits through pleiotropy analysis.

In our analysis, the UK Biobank Europeans was a part of data for identifying BP variants and pleiotropic variants. We then constructed the cGRS and pGRS in UK Biobank data by using the estimated effect sizes of the independent genome wide significant variants from UKB-ICBP summary statistics as the weights. This procedure was used in Evangelou et al.^6^ and was not involved in a model selection. However, there is a potential winner’s curse effect as suggested by Evangelou et al. When we applied the cGRS and pGRS to the UK Biobank Africans and Asians, we again observed the improved R^2^ (Supplementary Table 9), suggesting that adding cGRS improves the prediction power.

Our analysis avoided an examination of the interaction of individual variants and age because of insufficient power. We were able to observe interaction effects of both cGRS and pGRS with age in UKB Europeans for SBP, DBP, PP and HTN although the interaction for CVD was only significant for pGRS (**Table 2**). Age-pGRS interactions were also replicated in Asians despite a substantially smaller sample size. We observed that the interaction contribution to phenotype variation was consistently small (0.1% to 0.3% BP heritability in both UKB Europeans and Asians). The negative interaction contributions of both cGRS and pGRS to DBP may partially explain the decline of DBP after 60 years older ^48^. In comparison, the interaction of age and cGRS was positive for SBP, suggesting genetic effects on SBP increases in older individuals. As noted, the cGRS interaction effects in UKB Europeans could not be observed in UKB Africans or Asians, likely as a result of the latter’s smaller sample size. In comparison, the age-modulated interaction of pGRS was observed in both UKB European and Asian subjects, indicating stronger pGRS interactions than for the GRS. In our functional annotation analysis, we observed a wider range of BP related tissues and biological pathways for the BP pleiotropic variants than mediation variants, which implies that pleiotropic variants are influenced by a wider range of environmental factors and therefore continue to make genetic contributions over the life span.

Our analysis approach bears some similarity with a recent developed GWAS-by-subtraction^49^. However, there are significant differences. 1) The GWAS-by-subtraction is based on a genomic structure equation model (genomic-SEM) and our method is based on MR. 2) GWAS-by-subtraction assumes that all genetic effects on one trait affect the other. In contrast, our method does not make this assumption. Instead, our method estimates the causal effects in both directions. 3) The GWAS-by-subtraction tests the null hypothesis 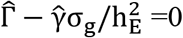, where 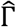 and 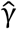 are the estimated effect sizes of a SNP on two traits (trait 1 and 2), σ_g_ is the genetic covariance between the two traits and 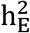 is the heritability of trait 2. In contrast, our method tests the null hypothesis 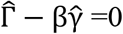 and 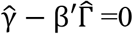 where β and β′ are the two causal effects estimated from MR. Noted that 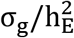 is not the same as causal effect β when pleiotropic variants exist. The GWAS-by-subtraction includes the pleiotropic variants in estimating genetic covariance and our method does not. Therefore, our method is less affected by pleiotropic variants. However, further research is warranted in understanding these two methods better.

Our analysis was performed on two highly correlated BP traits, which led to define mediation and pleiotropy variants. We were able to identify a “core” mediation pathway shared by both SBP and DBP, and a pleiotropy pathway that has different effects to SBP and DBP. Our method is therefore useful in analyzing different symptoms of a disease for understanding the biological mechanism of the disease. Furthermore, our approach can also be applied to less genetically correlated phenotypes to identify pleiotropic variants. In this case, we will less likely to identify “core” phenotypes as we observed in BP traits.

Our study also supports an omnigenic model for complex traits^50-52^. In fact, it could be inferred that pleiotropic variants act on multiple peripheral genes to impact the expression of core genes. As a result, pleiotropic variants have weak effects on a phenotype and are more difficult to detect in a traditional BP GWAS that focuses on single trait analysis, as observed here. In comparison, mediation variants may be more likely to occur in core genes. We acknowledge that the data presented here can only provide suggestive rather than conclusive evidence for that hypothesis.

In conclusion, our new findings here include identifying 815 independent BP pleiotropic variants - 201 of which were not previously identified in BP GWAS in the UKB-ICBP study; of these, 118 were confirmed including 18 novel BP loci. In addition, 219 novel BP signals and 40 novel loci were identified by combining UKB-ICBP and MVP. Our way to construct polygenic risk score represents a substantial advance in understanding the genetic architecture of the highly correlated SBP and DBP.

## Data Availability

Full summary statistics related to UKB-ICBP were obtained through request to the authors of UKB-ICBP Paul Elliott or Mark Caulfield. Summary statistics relating to the Million Veteran Program (MVP) are publically available with the accession code phs001672.v1.p1: https://www.ncbi.nlm.nih.gov/projects/gap/cgi-bin/study.cgi?study_id=phs001672.v1.p1. The UK BioBank data are available upon application to the UKBiobank (https://www.ukbiobank.ac.uk). The coronary artery disease and myocardial infraction summary statistics can be downloaded at http://www.cardiogramplusc4d.org/data-downloads/; Stroke summary statistics can be downloaded at: http://megastroke.org/privacy.html.

## URLs

CTG-View: https://view.genoma.io/

Depict: https://data.broadinstitute.org/mpg/depict/

GTEx: www.gtexportal.org

FUMA: https://fuma.ctglab.nl/

IMRP: https://github.com/XiaofengZhuCase/IMRP

MRmix://github.com/gqi/MRMix

## Acknowledgements

This work was supported by grant HG011052 (to XZ) from the National Human Genome Research Institute (NHGRI) and HL086694 (A.C.) from the National Heart, Lung and Blood Institute. We thank Dr. Jacklyn Hellwege for providing the list of published BP loci. Data on coronary artery disease / myocardial infarction have been contributed by CARDIoGRAMplusC4D investigators and have been downloaded from www.CARDIOGRAMPLUSC4D.ORG’. Data on coronary artery disease / myocardial infarction have been contributed by the Myocardial Infarction Genetics and CARDIoGRAM Exome investigators and have been downloaded from www.CARDIOGRAMPLUSC4D.ORG The MEGASTROKE project received funding from sources specified at http://www.megastroke.org/acknowledgments.html.

## Author contributions

X.Z conceived and designed the study. X.Z, L.Z. and H.W. performed analysis. X.Z. drafted the initial manuscript and A.C. edited the manuscript.. All authors critically revised and approved the manuscript.

## Supplementary Materials

**Supplementary Figure 1.**
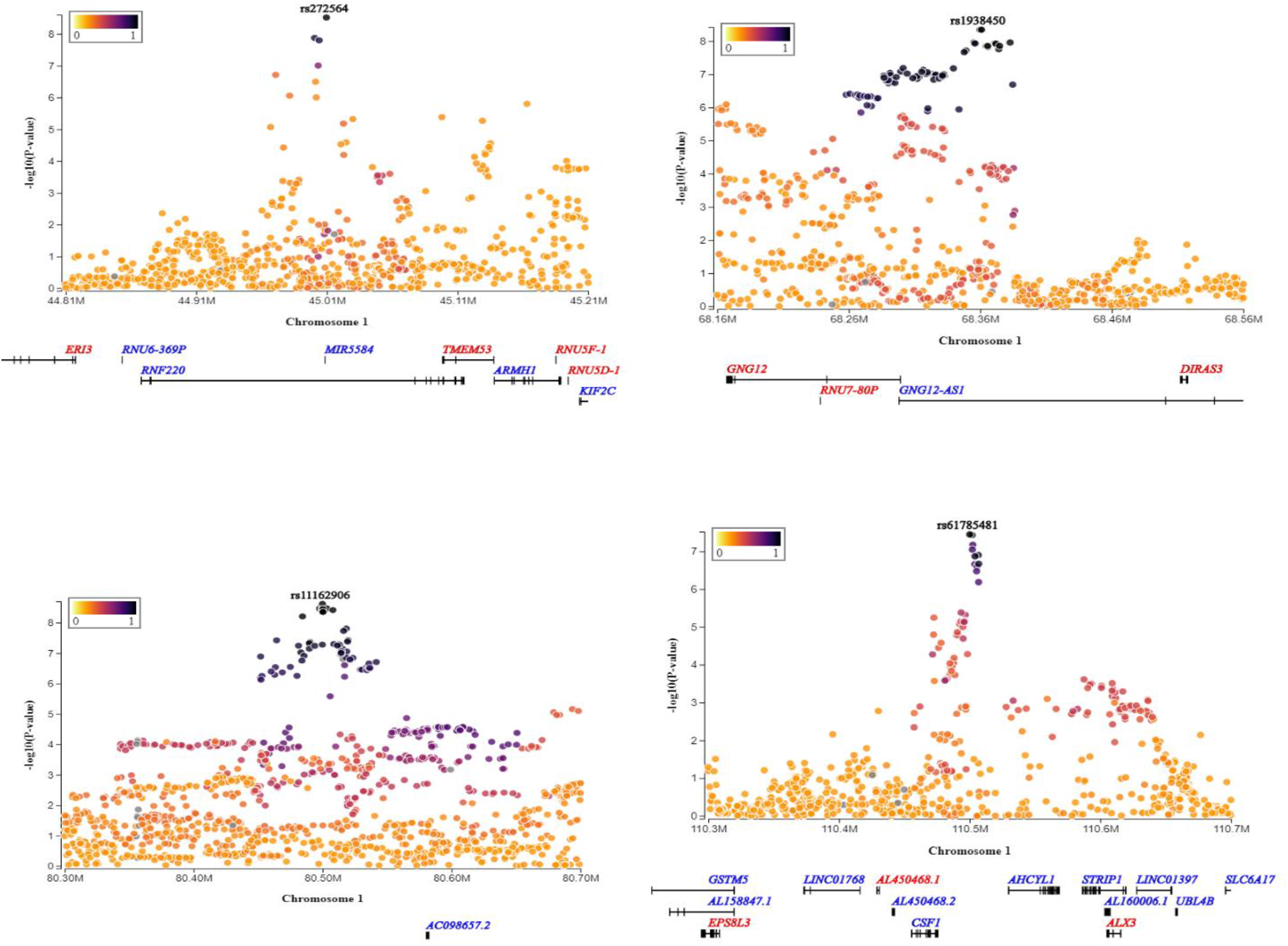
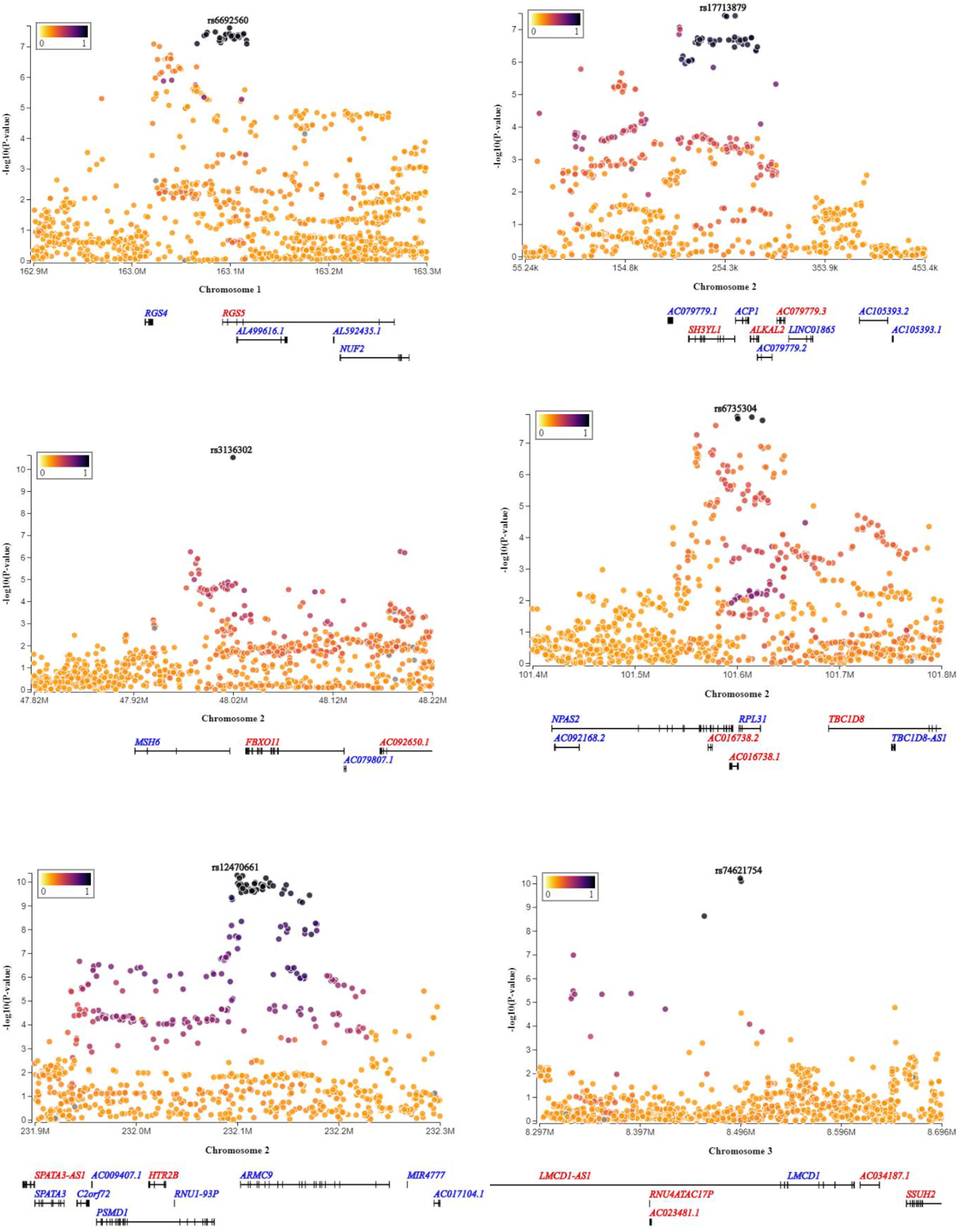
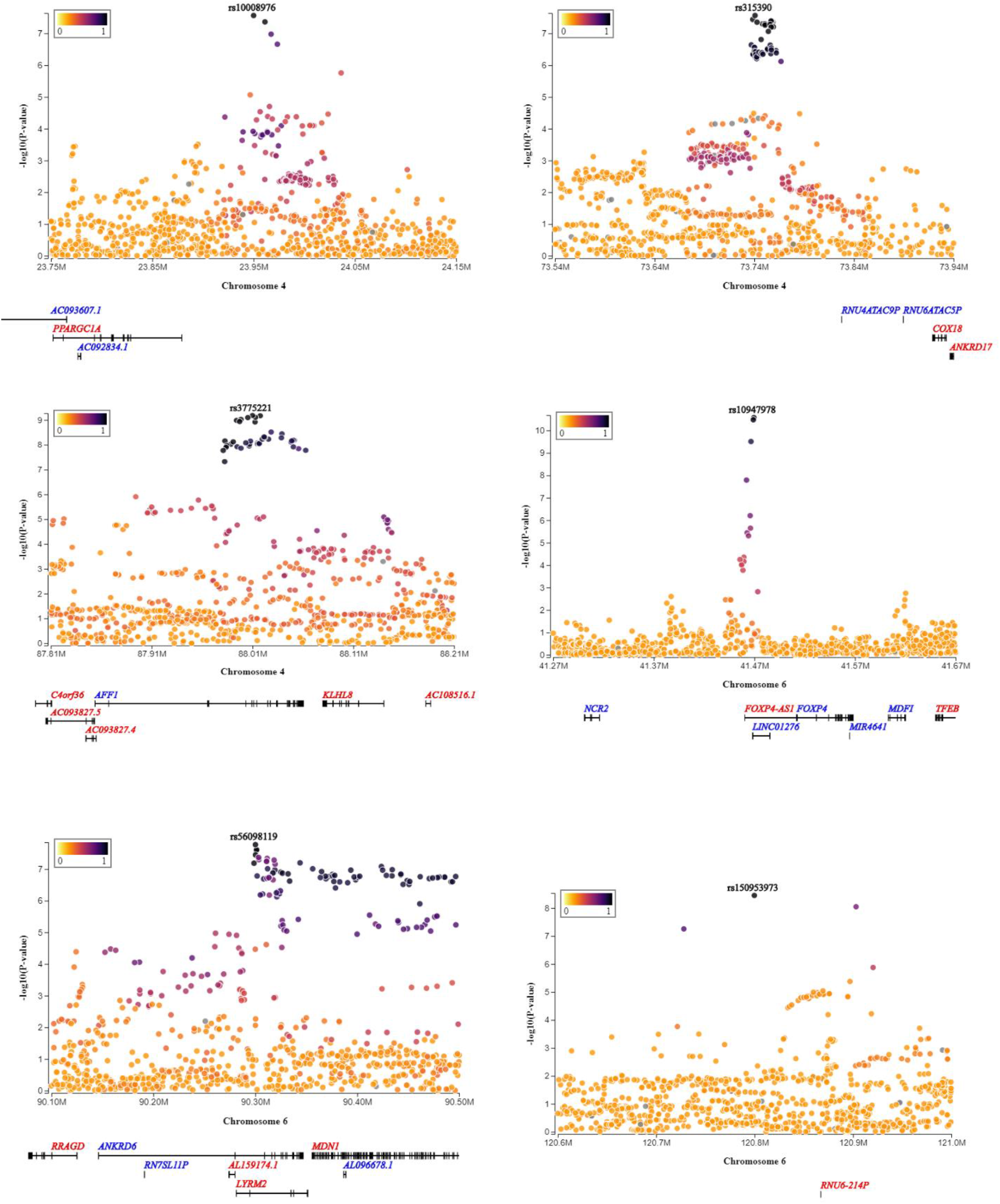
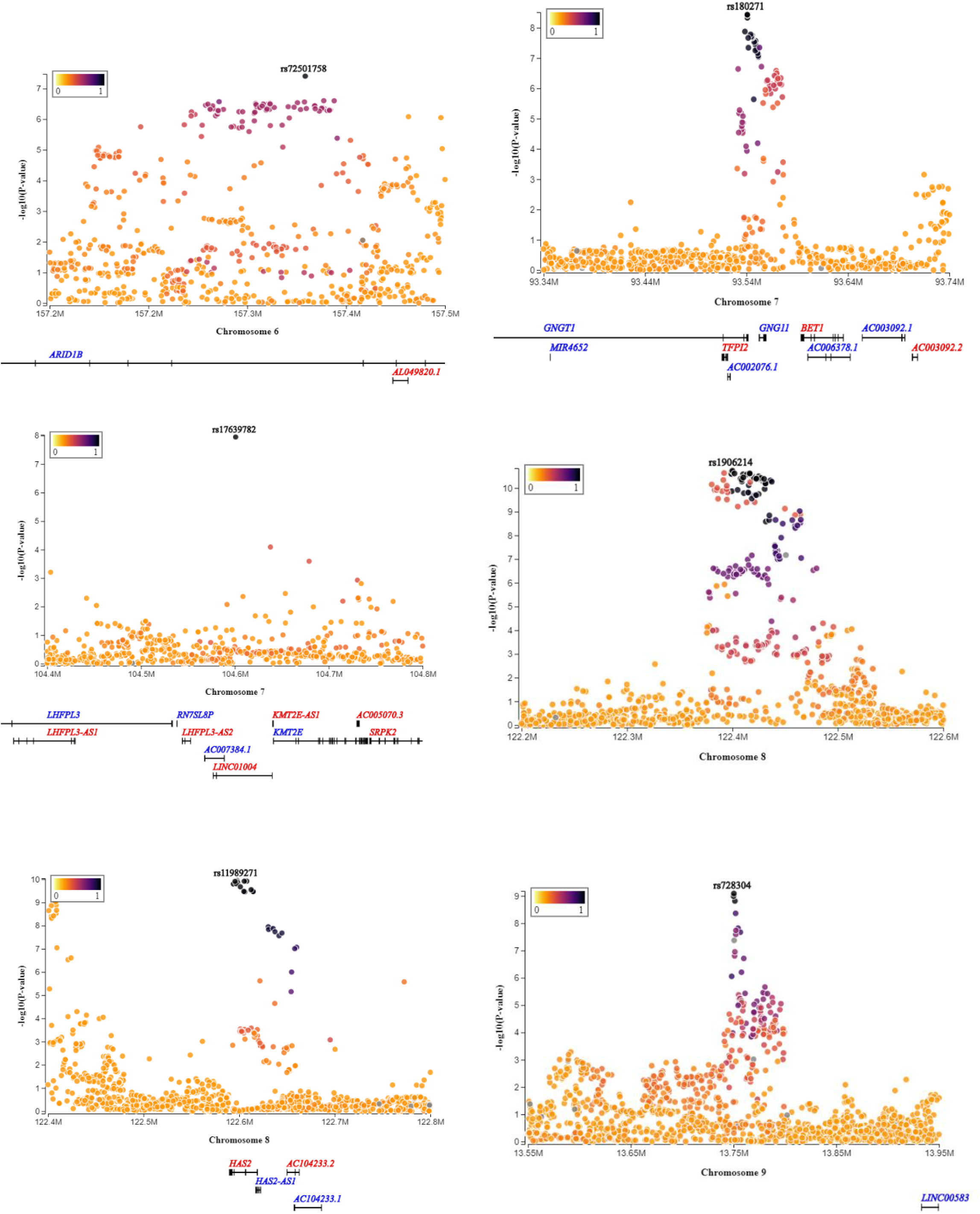
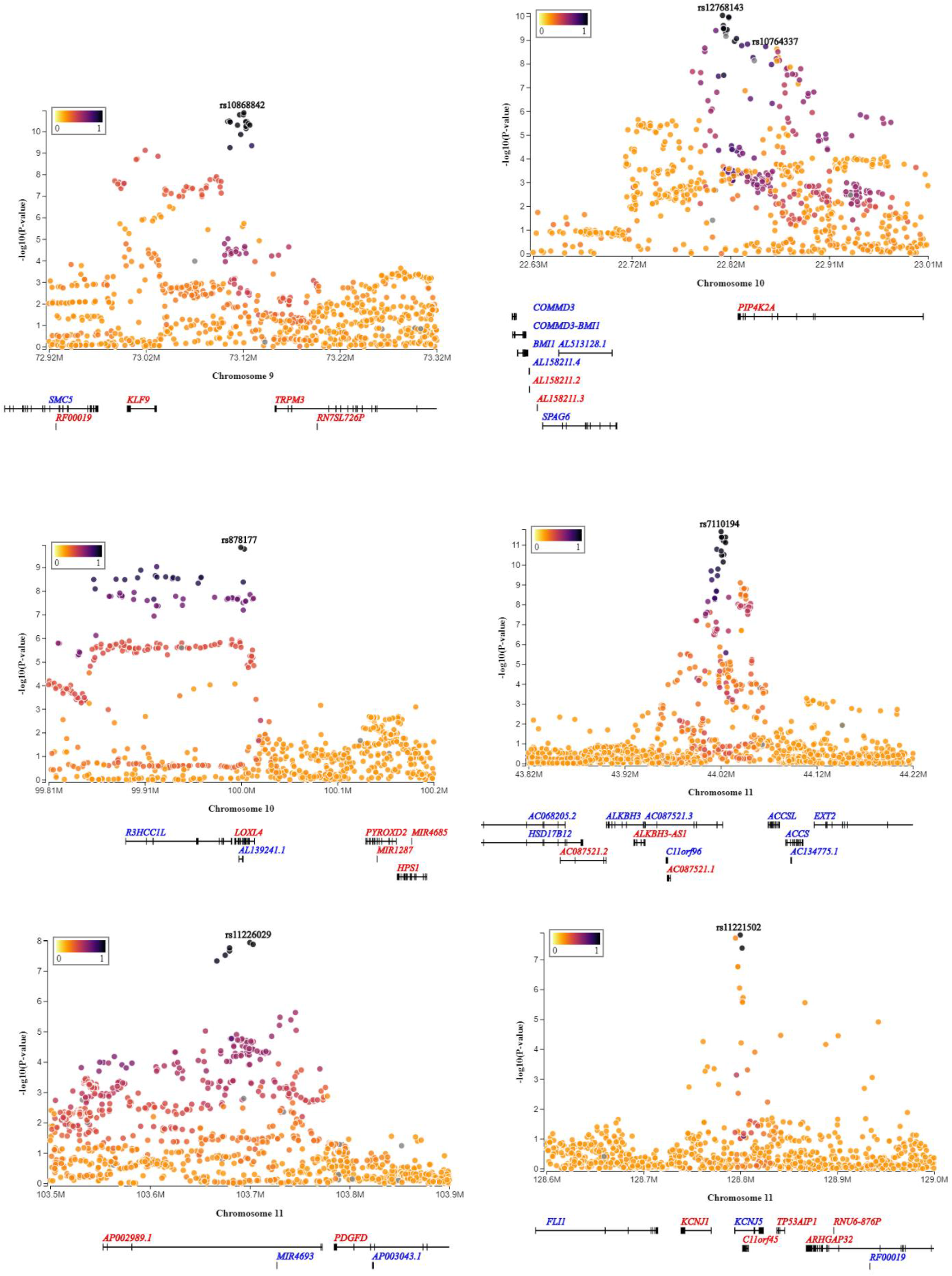
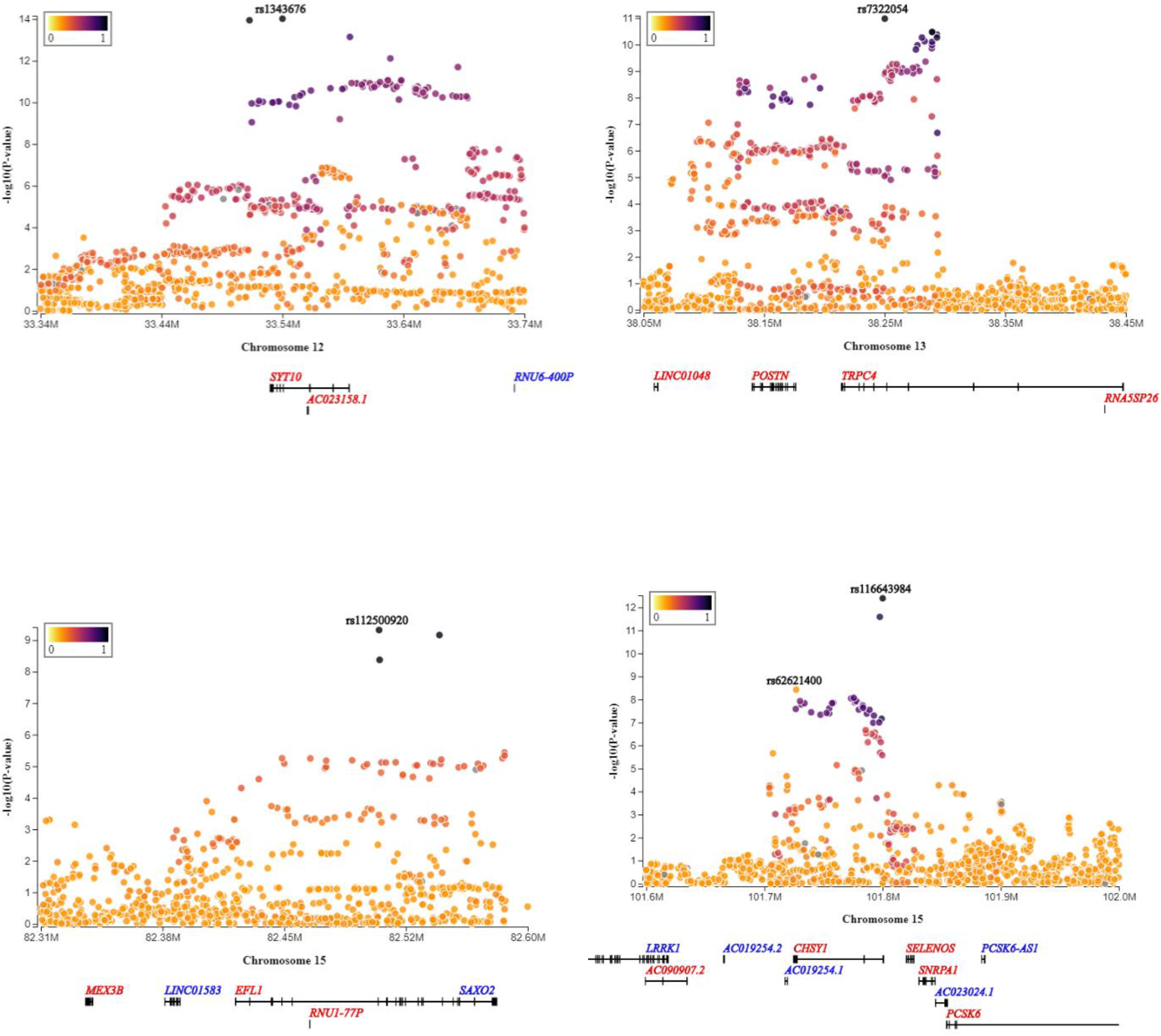
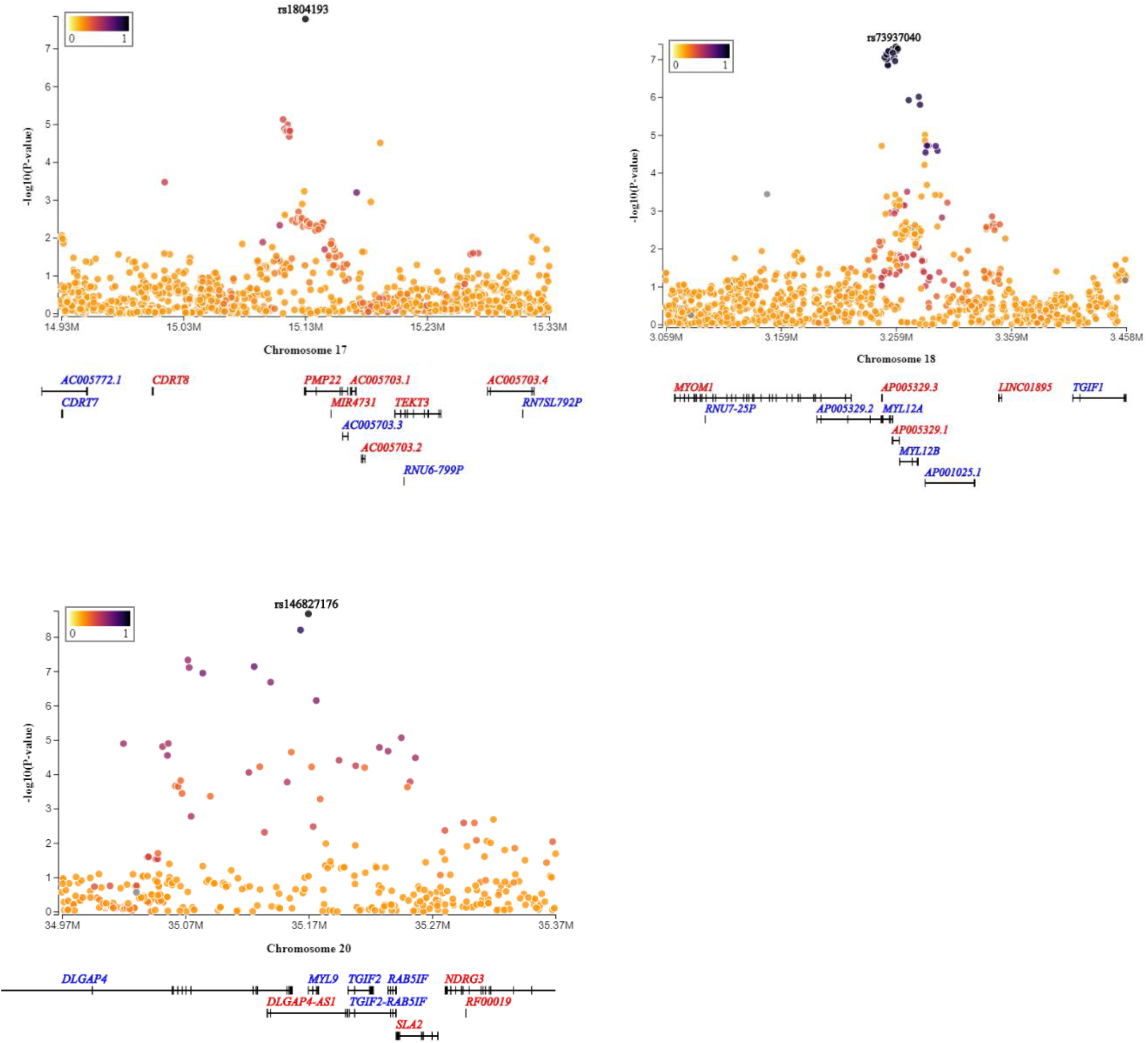
Locus zoom plots for 35 novel loci (selected from Table 1)

**Supplementary Figure2. A.**
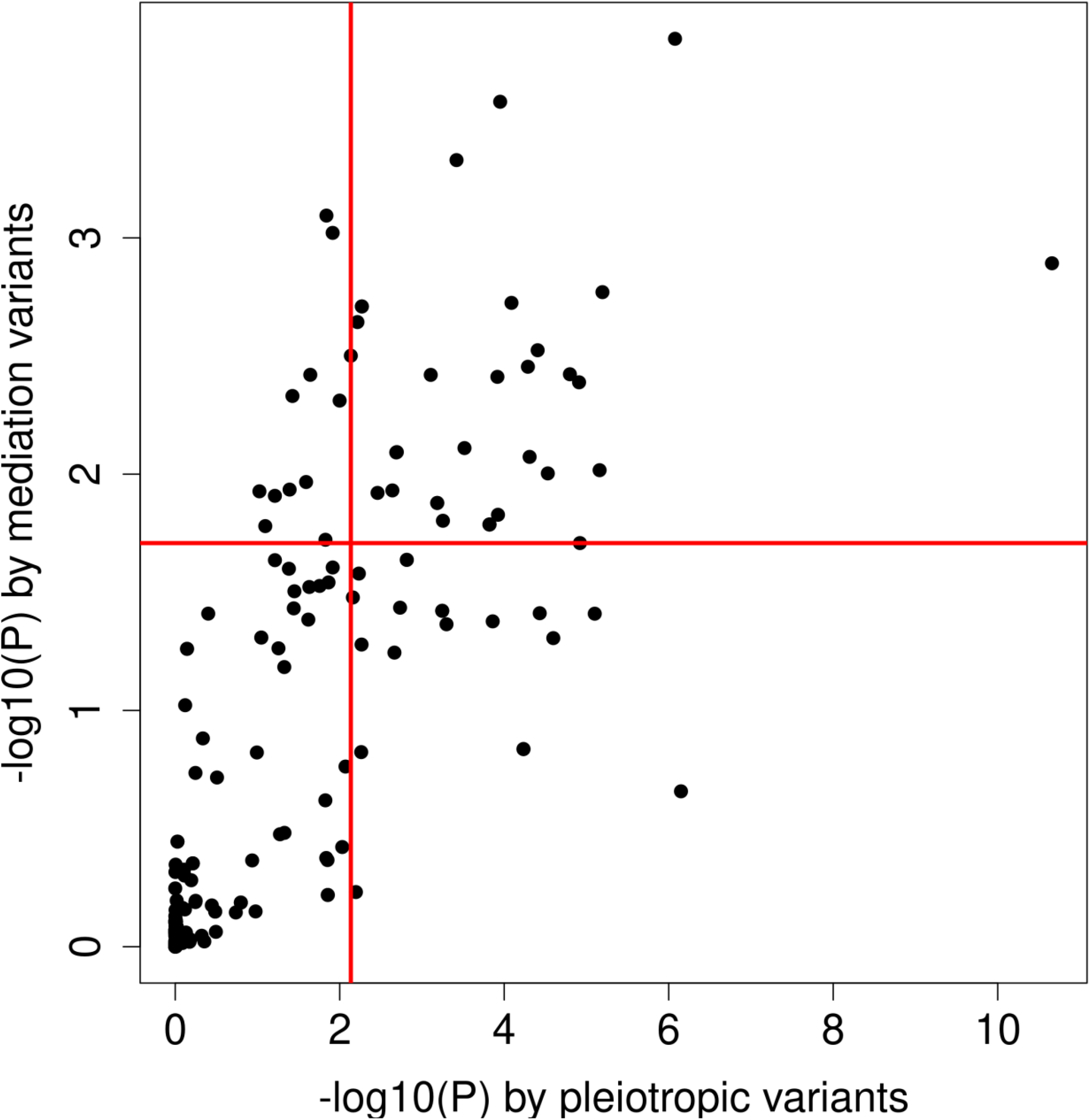
Tissue enrichment correlation using mediation and pleiotropic variants. Each dot represents a tissue. Above the red horizontal line are the tissues enriched by mediation evidence with FDR<5%. The points falling in the right side of the red vertical line are the tissues enriched by pleiotropy. (correlation=0.78)

**Supplementary Figure2. B.**
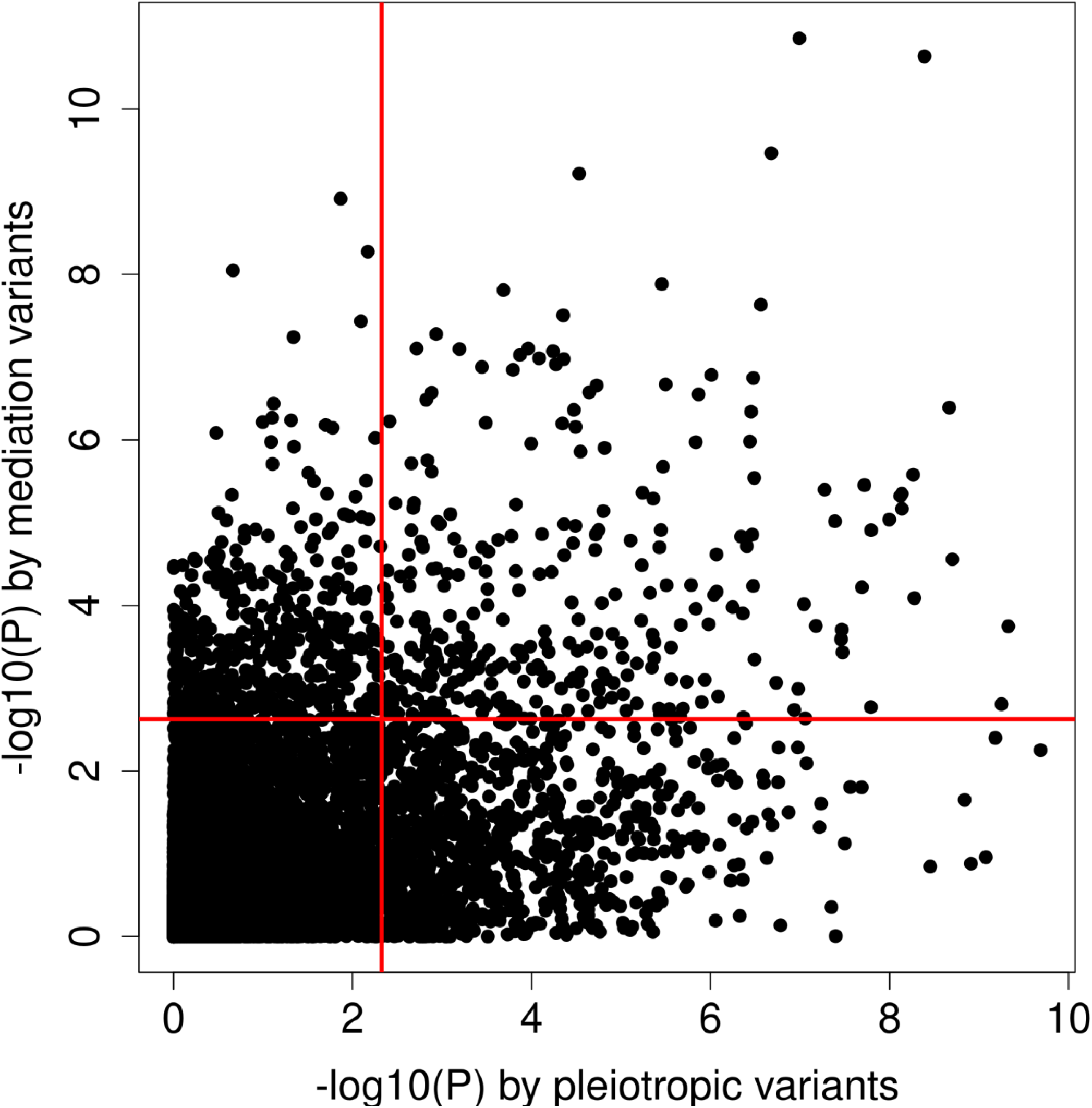
Pathway enrichment correlation using mediation and pleiotropic variants. Each dot represents a gene set pathway. Above the red horizontal line are the pathways enriched by mediation evidence with FDR<5%. The points falling in the right side of the red vertical line are the pathways enriched by pleiotropy evidence. (Spearman Rank correlation=0.552, -log10(P) Pearson correlation=0.478)

## MEGASTROKE CONSORTIUM

Rainer Malik ^1^, Ganesh Chauhan ^2^, Matthew Traylor ^3^, Muralidharan Sargurupremraj ^4,5^, Yukinori Okada ^6,7,8^, Aniket Mishra ^4,5^, Loes Rutten-Jacobs ^3^, Anne-Katrin Giese ^9^, Sander W van der Laan ^10^, Solveig Gretarsdottir ^11^, Christopher D Anderson ^12,13,14,14^, Michael Chong ^15^, Hieab HH Adams ^16,17^, Tetsuro Ago ^18^, Peter Almgren ^19^, Philippe Amouyel ^20,21^, Hakan Ay ^22,13^, Traci M Bartz ^23^, Oscar R Benavente ^24^, Steve Bevan ^25^, Giorgio B Boncoraglio ^26^, Robert D Brown, Jr. ^27^, Adam S Butterworth ^28,29^, Caty Carrera ^30,31^, Cara L Carty ^32,33^, Daniel I Chasman ^34,35^, Wei-Min Chen ^36^, John W Cole ^37^, Adolfo Correa ^38^, Ioana Cotlarciuc ^39^, Carlos Cruchaga ^40,41^, John Danesh ^28,42,43,44^, Paul IW de Bakker ^45,46^, Anita L DeStefano ^47,48^, Marcel den Hoed ^49^, Qing Duan ^50^, Stefan T Engelter ^51,52^, Guido J Falcone ^53,54^, Rebecca F Gottesman ^55^, Raji P Grewal ^56^, Vilmundur Gudnason ^57,58^, Stefan Gustafsson ^59^, Jeffrey Haessler ^60^, Tamara B Harris ^61^, Ahamad Hassan ^62^, Aki S Havulinna ^63,64^, Susan R Heckbert ^65^, Elizabeth G Holliday ^66,67^, George Howard ^68^, Fang-Chi Hsu ^69^, Hyacinth I Hyacinth ^70^, M Arfan Ikram ^16^, Erik Ingelsson ^71,72^, Marguerite R Irvin ^73^, Xueqiu Jian ^74^, Jordi Jiménez-Conde ^75^, Julie A Johnson ^76,77^, J Wouter Jukema ^78^, Masahiro Kanai ^6,7,79^, Keith L Keene ^80,81^, Brett M Kissela ^82^, Dawn O Kleindorfer ^82^, Charles Kooperberg ^60^, Michiaki Kubo ^83^, Leslie A Lange ^84^, Carl D Langefeld ^85^, Claudia Langenberg ^86^, Lenore J Launer ^87^, Jin-Moo Lee ^88^, Robin Lemmens ^89,90^, Didier Leys ^91^, Cathryn M Lewis ^92,93^, Wei-Yu Lin ^28,94^, Arne G Lindgren ^95,96^, Erik Lorentzen ^97^, Patrik K Magnusson ^98^, Jane Maguire ^99^, Ani Manichaikul ^36^, Patrick F McArdle ^100^, James F Meschia ^101^, Braxton D Mitchell ^100,102^, Thomas H Mosley ^103,104^, Michael A Nalls ^105,106^, Toshiharu Ninomiya ^107^, Martin J O’Donnell ^15,108^, Bruce M Psaty ^109,110,111,112^, Sara L Pulit ^113,45^, Kristiina Rannikmäe ^114,115^, Alexander P Reiner ^65,116^, Kathryn M Rexrode ^117^, Kenneth Rice ^118^, Stephen S Rich ^36^, Paul M Ridker ^34,35^, Natalia S Rost ^9,13^, Peter M Rothwell ^119^, Jerome I Rotter ^120,121^, Tatjana Rundek ^122^, Ralph L Sacco ^122^, Saori Sakaue ^7,123^, Michele M Sale ^124^, Veikko Salomaa ^63^, Bishwa R Sapkota ^125^, Reinhold Schmidt ^126^, Carsten O Schmidt ^127^, Ulf Schminke ^128^, Pankaj Sharma ^39^, Agnieszka Slowik ^129^, Cathie LM Sudlow ^114,115^, Christian Tanislav ^130^, Turgut Tatlisumak ^131,132^, Kent D Taylor ^120,121^, Vincent NS Thijs ^133,134^, Gudmar Thorleifsson ^11^, Unnur Thorsteinsdottir ^11^, Steffen Tiedt ^1^, Stella Trompet ^135^, Christophe Tzourio ^5,136,137^, Cornelia M van Duijn ^138,139^, Matthew Walters ^140^, Nicholas J Wareham ^86^, Sylvia Wassertheil-Smoller ^141^, James G Wilson ^142^, Kerri L Wiggins ^109^, Qiong Yang ^47^, Salim Yusuf ^15^, Najaf Amin ^16^, Hugo S Aparicio ^185,48^, Donna K Arnett ^186^, John Attia ^187^, Alexa S Beiser ^47,48^, Claudine Berr ^188^, Julie E Buring ^34,35^, Mariana Bustamante ^189^, Valeria Caso ^190^, Yu-Ching Cheng ^191^, Seung Hoan Choi ^192,48^, Ayesha Chowhan ^185,48^, Natalia Cullell ^31^, Jean-François Dartigues ^193,194^, Hossein Delavaran ^95,96^, Pilar Delgado ^195^, Marcus Dörr ^196,197^, Gunnar Engström ^19^, Ian Ford ^198^, Wander S Gurpreet ^199^, Anders Hamsten ^200,201^, Laura Heitsch ^202^, Atsushi Hozawa ^203^, Laura Ibanez ^204^, Andreea Ilinca ^95,96^, Martin Ingelsson ^205^, Motoki Iwasaki ^206^, Rebecca D Jackson ^207^, Katarina Jood ^208^, Pekka Jousilahti ^63^, Sara Kaffashian ^4,5^, Lalit Kalra ^209^, Masahiro Kamouchi ^210^, Takanari Kitazono ^211^, Olafur Kjartansson ^212^, Manja Kloss ^213^, Peter J Koudstaal ^214^, Jerzy Krupinski ^215^, Daniel L Labovitz ^216^, Cathy C Laurie ^118^, Christopher R Levi ^217^, Linxin Li ^218^, Lars Lind ^219^, Cecilia M Lindgren ^220,221^, Vasileios Lioutas ^222,48^, Yong Mei Liu ^223^, Oscar L Lopez ^224^, Hirata Makoto ^225^, Nicolas Martinez-Majander ^172^, Koichi Matsuda ^225^, Naoko Minegishi ^203^, Joan Montaner ^226^, Andrew P Morris ^227,228^, Elena Muiño ^31^, Martina Müller-Nurasyid ^229,230,231^, Bo Norrving ^95,96^, Soichi Ogishima ^203^, Eugenio A Parati ^232^, Leema Reddy Peddareddygari ^56^, Nancy L Pedersen ^98,233^, Joanna Pera ^129^, Markus Perola ^63,234^, Alessandro Pezzini ^235^, Silvana Pileggi ^236^, Raquel Rabionet ^237^, Iolanda Riba-Llena ^30^, Marta Ribasés ^238^, Jose R Romero ^185,48^, Jaume Roquer ^239,240^, Anthony G Rudd ^241,242^, Antti-Pekka Sarin ^243,244^, Ralhan Sarju ^199^, Chloe Sarnowski ^47,48^, Makoto Sasaki ^245^, Claudia L Satizabal ^185,48^, Mamoru Satoh ^245^, Naveed Sattar ^246^, Norie Sawada ^206^, Gerli Sibolt ^172^, Ásgeir Sigurdsson ^247^, Albert Smith ^248^, Kenji Sobue ^245^, Carolina Soriano-Tárraga ^240^, Tara Stanne ^249^, O Colin Stine ^250^, David J Stott ^251^, Konstantin Strauch ^229,252^, Takako Takai ^203^, Hideo Tanaka ^253,254^, Kozo Tanno ^245^, Alexander Teumer ^255^, Liisa Tomppo ^172^, Nuria P Torres-Aguila ^31^, Emmanuel Touze ^256,257^, Shoichiro Tsugane ^206^, Andre G Uitterlinden ^258^, Einar M Valdimarsson ^259^, Sven J van der Lee ^16^, Henry Völzke ^255^, Kenji Wakai ^253^, David Weir ^260^, Stephen R Williams ^261^, Charles DA Wolfe ^241,242^, Quenna Wong ^118^, Huichun Xu ^191^, Taiki Yamaji ^206^, Dharambir K Sanghera ^125,169,170^, Olle Melander ^19^, Christina Jern ^171^, Daniel Strbian ^172,173^, Israel Fernandez-Cadenas ^31,30^, W T Longstreth, Jr ^174,65^, Arndt Rolfs ^175^, Jun Hata ^107^, Daniel Woo ^82^, Jonathan Rosand ^12,13,14^, Guillaume Pare ^15^, Jemma C Hopewell ^176^, Danish Saleheen ^177^, Kari Stefansson ^11,178^, Bradford B Worrall ^179^, Steven J Kittner ^37^, Sudha Seshadri ^180,48^, Myriam Fornage ^74,181^, Hugh S Markus ^3^, Joanna MM Howson ^28^, Yoichiro Kamatani ^6,182^, Stephanie Debette ^4,5^, Martin Dichgans ^1,183,184^

1 Institute for Stroke and Dementia Research (ISD), University Hospital, LMU Munich, Munich, Germany

2 Centre for Brain Research, Indian Institute of Science, Bangalore, India

3 Stroke Research Group, Division of Clinical Neurosciences, University of Cambridge, UK

4 INSERM U1219 Bordeaux Population Health Research Center, Bordeaux, France

5 University of Bordeaux, Bordeaux, France

6 Laboratory for Statistical Analysis, RIKEN Center for Integrative Medical Sciences, Yokohama, Japan

7 Department of Statistical Genetics, Osaka University Graduate School of Medicine, Osaka, Japan

8 Laboratory of Statistical Immunology, Immunology Frontier Research Center (WPI-IFReC), Osaka University, Suita, Japan.

9 Department of Neurology, Massachusetts General Hospital, Harvard Medical School, Boston, MA, USA

10 Laboratory of Experimental Cardiology, Division of Heart and Lungs, University Medical Center Utrecht, University of Utrecht, Utrecht,Netherlands

11 deCODE genetics/AMGEN inc, Reykjavik, Iceland

12 Center for Genomic Medicine, Massachusetts General Hospital (MGH), Boston, MA, USA

13 J. Philip Kistler Stroke Research Center, Department of Neurology, MGH, Boston, MA, USA

14 Program in Medical and Population Genetics, Broad Institute, Cambridge, MA, USA

15 Population Health Research Institute, McMaster University, Hamilton, Canada

16 Department of Epidemiology, Erasmus University Medical Center, Rotterdam, Netherlands

17 Department of Radiology and Nuclear Medicine, Erasmus University Medical Center, Rotterdam, Netherlands

18 Department of Medicine and Clinical Science, Graduate School of Medical Sciences, Kyushu University, Fukuoka, Japan

19 Department of Clinical Sciences, Lund University, Malmö, Sweden

20 Univ. Lille, Inserm, Institut Pasteur de Lille, LabEx DISTALZ-UMR1167, Risk factors and molecular determinants of aging-related diseases, F-59000 Lille, France

21 Centre Hosp. Univ Lille, Epidemiology and Public Health Department, F-59000 Lille, France

22 AA Martinos Center for Biomedical Imaging, Department of Radiology, Massachusetts General Hospital, Harvard Medical School, Boston, MA, USA

23 Cardiovascular Health Research Unit, Departments of Biostatistics and Medicine, University of Washington, Seattle, WA, USA

24 Division of Neurology, Faculty of Medicine, Brain Research Center, University of British Columbia, Vancouver, Canada

25 School of Life Science, University of Lincoln, Lincoln, UK

26 Department of Cerebrovascular Diseases, Fondazione IRCCS Istituto Neurologico “Carlo Besta”, Milano, Italy

27 Department of Neurology, Mayo Clinic Rochester, Rochester, MN, USA

28 MRC/BHF Cardiovascular Epidemiology Unit, Department of Public Health and Primary Care, University of Cambridge, Cambridge, UK

29 The National Institute for Health Research Blood and Transplant Research Unit in Donor Health and Genomics, University of Cambridge, UK

30 Neurovascular Research Laboratory, Vall d’Hebron Institut of Research, Neurology and Medicine Departments-Universitat Autònoma de Barcelona, Vall d’Hebrón Hospital, Barcelona, Spain

31 Stroke Pharmacogenomics and Genetics, Fundacio Docència i Recerca MutuaTerrassa, Terrassa, Spain

32 Children’s Research Institute, Children’s National Medical Center, Washington, DC, USA

33 Center for Translational Science, George Washington University, Washington, DC, USA

34 Division of Preventive Medicine, Brigham and Women’s Hospital, Boston, MA, USA

35 Harvard Medical School, Boston, MA, USA

36 Center for Public Health Genomics, Department of Public Health Sciences, University of Virginia, Charlottesville, VA, USA

37 Department of Neurology, University of Maryland School of Medicine and Baltimore VAMC, Baltimore, MD, USA

38 Departments of Medicine, Pediatrics and Population Health Science, University of Mississippi Medical Center, Jackson, MS, USA

39 Institute of Cardiovascular Research, Royal Holloway University of London, UK & Ashford and St Peters Hospital, Surrey UK

40 Department of Psychiatry,The Hope Center Program on Protein Aggregation and Neurodegeneration (HPAN),Washington University, School of Medicine, St. Louis, MO, USA

41 Department of Developmental Biology, Washington University School of Medicine, St. Louis, MO, USA

42 NIHR Blood and Transplant Research Unit in Donor Health and Genomics, Department of Public Health and Primary Care, University of Cambridge, Cambridge, UK

43 Wellcome Trust Sanger Institute, Wellcome Trust Genome Campus, Hinxton, Cambridge, UK

44 British Heart Foundation, Cambridge Centre of Excellence, Department of Medicine, University of Cambridge, Cambridge, UK

45 Department of Medical Genetics, University Medical Center Utrecht, Utrecht, Netherlands

46 Department of Epidemiology, Julius Center for Health Sciences and Primary Care, University Medical Center Utrecht, Utrecht, Netherlands

47 Boston University School of Public Health, Boston, MA, USA

48 Framingham Heart Study, Framingham, MA, USA

49 Department of Immunology, Genetics and Pathology and Science for Life Laboratory, Uppsala University, Uppsala, Sweden

50 Department of Genetics, University of North Carolina, Chapel Hill, NC, USA

51 Department of Neurology and Stroke Center, Basel University Hospital, Switzerland

52 Neurorehabilitation Unit, University and University Center for Medicine of Aging and Rehabilitation Basel, Felix Platter Hospital, Basel, Switzerland

53 Department of Neurology, Yale University School of Medicine, New Haven, CT, USA

54 Program in Medical and Population Genetics, The Broad Institute of Harvard and MIT, Cambridge, MA, USA

55 Department of Neurology, Johns Hopkins University School of Medicine, Baltimore, MD, USA

56 Neuroscience Institute, SF Medical Center, Trenton, NJ, USA

57 Icelandic Heart Association Research Institute, Kopavogur, Iceland

58 University of Iceland, Faculty of Medicine, Reykjavik, Iceland

59 Department of Medical Sciences, Molecular Epidemiology and Science for Life Laboratory, Uppsala University, Uppsala, Sweden

60 Division of Public Health Sciences, Fred Hutchinson Cancer Research Center, Seattle, WA, USA

61 Laboratory of Epidemiology and Population Science, National Institute on Aging, National Institutes of Health, Bethesda, MD, USA

62 Department of Neurology, Leeds General Infirmary, Leeds Teaching Hospitals NHS Trust, Leeds, UK

63 National Institute for Health and Welfare, Helsinki, Finland

64 FIMM - Institute for Molecular Medicine Finland, Helsinki, Finland

65 Department of Epidemiology, University of Washington, Seattle, WA, USA

66 Public Health Stream, Hunter Medical Research Institute, New Lambton, Australia

67 Faculty of Health and Medicine, University of Newcastle, Newcastle, Australia

68 School of Public Health, University of Alabama at Birmingham, Birmingham, AL, USA

69 Department of Biostatistical Sciences, Wake Forest School of Medicine, Winston-Salem, NC, USA

70 Aflac Cancer and Blood Disorder Center, Department of Pediatrics, Emory University School of Medicine, Atlanta, GA, USA

71 Department of Medicine, Division of Cardiovascular Medicine, Stanford University School of Medicine, CA, USA

72 Department of Medical Sciences, Molecular Epidemiology and Science for Life Laboratory, Uppsala University, Uppsala, Sweden

73 Epidemiology, School of Public Health, University of Alabama at Birmingham, USA

74 Brown Foundation Institute of Molecular Medicine, University of Texas Health Science Center at Houston, Houston, TX, USA

75 Neurovascular Research Group (NEUVAS), Neurology Department, Institut Hospital del Mar d’Investigació Mèdica, Universitat Autònoma de Barcelona, Barcelona, Spain

76 Department of Pharmacotherapy and Translational Research and Center for Pharmacogenomics, University of Florida, College of Pharmacy, Gainesville, FL, USA

77 Division of Cardiovascular Medicine, College of Medicine, University of Florida, Gainesville, FL, USA

78 Department of Cardiology, Leiden University Medical Center, Leiden, the Netherlands

79 Program in Bioinformatics and Integrative Genomics, Harvard Medical School, Boston, MA, USA

80 Department of Biology, East Carolina University, Greenville, NC, USA

81 Center for Health Disparities, East Carolina University, Greenville, NC, USA

82 University of Cincinnati College of Medicine, Cincinnati, OH, USA

83 RIKEN Center for Integrative Medical Sciences, Yokohama, Japan

84 Department of Medicine, University of Colorado Denver, Anschutz Medical Campus, Aurora, CO, USA

85 Center for Public Health Genomics and Department of Biostatistical Sciences, Wake Forest School of Medicine, Winston-Salem, NC, USA

86 MRC Epidemiology Unit, University of Cambridge School of Clinical Medicine, Institute of Metabolic Science, Cambridge Biomedical Campus, Cambridge, UK

87 Intramural Research Program, National Institute on Aging, National Institutes of Health, Bethesda, MD, USA

88 Department of Neurology, Radiology, and Biomedical Engineering, Washington University School of Medicine, St. Louis, MO, USA

89 KU Leuven – University of Leuven, Department of Neurosciences, Experimental Neurology, Leuven, Belgium

90 VIB Center for Brain & Disease Research, University Hospitals Leuven, Department of Neurology, Leuven, Belgium

91 Univ.-Lille, INSERM U 1171. CHU Lille. Lille, France

92 Department of Medical and Molecular Genetics, King’s College London, London, UK

93 SGDP Centre, Institute of Psychiatry, Psychology & Neuroscience, King’s College London, London, UK

94 Northern Institute for Cancer Research, Paul O’Gorman Building, Newcastle University, Newcastle, UK

95 Department of Clinical Sciences Lund, Neurology, Lund University, Lund, Sweden

96 Department of Neurology and Rehabilitation Medicine, Skåne University Hospital, Lund, Sweden

97 Bioinformatics Core Facility, University of Gothenburg, Gothenburg, Sweden

98 Department of Medical Epidemiology and Biostatistics, Karolinska Institutet, Stockholm, Sweden

99 University of Technology Sydney, Faculty of Health, Ultimo, Australia

100 Department of Medicine, University of Maryland School of Medicine, MD, USA

101 Department of Neurology, Mayo Clinic, Jacksonville, FL, USA

102 Geriatrics Research and Education Clinical Center, Baltimore Veterans Administration Medical Center, Baltimore, MD, USA

103 Division of Geriatrics, School of Medicine, University of Mississippi Medical Center, Jackson, MS, USA

104 Memory Impairment and Neurodegenerative Dementia Center, University of Mississippi Medical Center, Jackson, MS, USA

105 Laboratory of Neurogenetics, National Institute on Aging, National institutes of Health, Bethesda, MD, USA

106 Data Tecnica International, Glen Echo MD, USA

107 Department of Epidemiology and Public Health, Graduate School of Medical Sciences, Kyushu University, Fukuoka, Japan

108 Clinical Research Facility, Department of Medicine, NUI Galway, Galway, Ireland

109 Cardiovascular Health Research Unit, Department of Medicine, University of Washington, Seattle, WA, USA

110 Department of Epidemiology, University of Washington, Seattle, WA

111 Department of Health Services, University of Washington, Seattle, WA, USA

112 Kaiser Permanente Washington Health Research Institute, Seattle, WA, USA

113 Brain Center Rudolf Magnus, Department of Neurology, University Medical Center Utrecht, Utrecht, The Netherlands

114 Usher Institute of Population Health Sciences and Informatics, University of Edinburgh, Edinburgh, UK

115 Centre for Clinical Brain Sciences, University of Edinburgh, Edinburgh, UK

116 Fred Hutchinson Cancer Research Center, University of Washington, Seattle, WA, USA

117 Department of Medicine, Brigham and Women’s Hospital, Boston, MA, USA

118 Department of Biostatistics, University of Washington, Seattle, WA, USA

119 Nuffield Department of Clinical Neurosciences, University of Oxford, UK

120 Institute for Translational Genomics and Population Sciences, Los Angeles Biomedical Research Institute at Harbor-UCLA Medical Center, Torrance, CA, USA

121 Division of Genomic Outcomes, Department of Pediatrics, Harbor-UCLA Medical Center, Torrance, CA, USA

122 Department of Neurology, Miller School of Medicine, University of Miami, Miami, FL, USA

123 Department of Allergy and Rheumatology, Graduate School of Medicine, the University of Tokyo, Tokyo, Japan

124 Center for Public Health Genomics, University of Virginia, Charlottesville, VA, USA

125 Department of Pediatrics, College of Medicine, University of Oklahoma Health Sciences Center, Oklahoma City, OK, USA

126 Department of Neurology, Medical University of Graz, Graz, Austria

127 University Medicine Greifswald, Institute for Community Medicine, SHIP-KEF, Greifswald, Germany

128 University Medicine Greifswald, Department of Neurology, Greifswald, Germany

129 Department of Neurology, Jagiellonian University, Krakow, Poland

130 Department of Neurology, Justus Liebig University, Giessen, Germany

131 Department of Clinical Neurosciences/Neurology, Institute of Neuroscience and Physiology, Sahlgrenska Academy at University of Gothenburg, Gothenburg, Sweden

132 Sahlgrenska University Hospital, Gothenburg, Sweden

133 Stroke Division, Florey Institute of Neuroscience and Mental Health, University of Melbourne, Heidelberg, Australia

134 Austin Health, Department of Neurology, Heidelberg, Australia

135 Department of Internal Medicine, Section Gerontology and Geriatrics, Leiden University Medical Center, Leiden, the Netherlands

136 INSERM U1219, Bordeaux, France

137 Department of Public Health, Bordeaux University Hospital, Bordeaux, France

138 Genetic Epidemiology Unit, Department of Epidemiology, Erasmus University Medical Center Rotterdam, Netherlands

139 Center for Medical Systems Biology, Leiden, Netherlands

140 School of Medicine, Dentistry and Nursing at the University of Glasgow, Glasgow, UK

141 Department of Epidemiology and Population Health, Albert Einstein College of Medicine, NY, USA

142 Department of Physiology and Biophysics, University of Mississippi Medical Center, Jackson, MS, USA

143 A full list of members and affiliations appears in the Supplementary Note

144 Department of Human Genetics, McGill University, Montreal, Canada

145 Department of Pathophysiology, Institute of Biomedicine and Translation Medicine, University of Tartu, Tartu, Estonia

146 Department of Cardiac Surgery, Tartu University Hospital, Tartu, Estonia

147 Clinical Gene Networks AB,Stockholm, Sweden

148 Department of Genetics and Genomic Sciences, The Icahn Institute for Genomics and Multiscale Biology Icahn School of Medicine at Mount Sinai, New York, NY, USA

149 Department of Pathophysiology, Institute of Biomedicine and Translation Medicine, University of Tartu, Biomeedikum, Tartu, Estonia

150 Integrated Cardio Metabolic Centre, Department of Medicine, Karolinska Institutet, Karolinska Universitetssjukhuset, Huddinge, Sweden.

151 Clinical Gene Networks AB, Stockholm, Sweden

152 Sorbonne Universités, UPMC Univ. Paris 06, INSERM, UMR_S 1166, Team Genomics & Pathophysiology of Cardiovascular Diseases, Paris, France

153 ICAN Institute for Cardiometabolism and Nutrition, Paris, France

154 Department of Biomedical Engineering, University of Virginia, Charlottesville, VA, USA

155 Group Health Research Institute, Group Health Cooperative, Seattle, WA, USA

156 Seattle Epidemiologic Research and Information Center, VA Office of Research and Development, Seattle, WA, USA

157 Cardiovascular Research Center, Massachusetts General Hospital, Boston, MA, USA

158 Department of Medical Research, Bærum Hospital, Vestre Viken Hospital Trust, Gjettum, Norway

159 Saw Swee Hock School of Public Health, National University of Singapore and National University Health System, Singapore

160 National Heart and Lung Institute, Imperial College London, London, UK

161 Department of Gene Diagnostics and Therapeutics, Research Institute, National Center for Global Health and Medicine, Tokyo, Japan

162 Department of Epidemiology, Tulane University School of Public Health and Tropical Medicine, New Orleans, LA, USA

163 Department of Cardiology,University Medical Center Groningen, University of Groningen, Netherlands

164 MRC-PHE Centre for Environment and Health, School of Public Health, Department of Epidemiology and Biostatistics, Imperial College London, London, UK

165 Department of Epidemiology and Biostatistics, Imperial College London, London, UK

166 Department of Cardiology, Ealing Hospital NHS Trust, Southall, UK

167 National Heart, Lung and Blood Research Institute, Division of Intramural Research, Population Sciences Branch, Framingham, MA, USA

168 A full list of members and affiliations appears at the end of the manuscript

169 Department of Phamaceutical Sciences, Collge of Pharmacy, University of Oklahoma Health Sciences Center, Oklahoma City, OK, USA

170 Oklahoma Center for Neuroscience, Oklahoma City, OK, USA

171 Department of Pathology and Genetics, Institute of Biomedicine, The Sahlgrenska Academy at University of Gothenburg, Gothenburg, Sweden

172 Department of Neurology, Helsinki University Hospital, Helsinki, Finland

173 Clinical Neurosciences, Neurology, University of Helsinki, Helsinki, Finland

174 Department of Neurology, University of Washington, Seattle, WA, USA

175 Albrecht Kossel Institute, University Clinic of Rostock, Rostock, Germany

176 Clinical Trial Service Unit and Epidemiological Studies Unit, Nuffield Department of Population Health, University of Oxford, Oxford, UK

177 Department of Genetics, Perelman School of Medicine, University of Pennsylvania, PA, USA

178 Faculty of Medicine, University of Iceland, Reykjavik, Iceland

179 Departments of Neurology and Public Health Sciences, University of Virginia School of Medicine, Charlottesville, VA, USA

180 Department of Neurology, Boston University School of Medicine, Boston, MA, USA

181 Human Genetics Center, University of Texas Health Science Center at Houston, Houston, TX, USA

182 Center for Genomic Medicine, Kyoto University Graduate School of Medicine, Kyoto, Japan

183 Munich Cluster for Systems Neurology (SyNergy), Munich, Germany

184 German Center for Neurodegenerative Diseases (DZNE), Munich, Germany

185 Boston University School of Medicine, Boston, MA, USA

186 University of Kentucky College of Public Health, Lexington, KY, USA

187 University of Newcastle and Hunter Medical Research Institute, New Lambton, Australia

188 Univ. Montpellier, Inserm, U1061, Montpellier, France

189 Centre for Research in Environmental Epidemiology, Barcelona, Spain

190 Department of Neurology, Università degli Studi di Perugia, Umbria, Italy

191 Department of Medicine, University of Maryland School of Medicine, Baltimore, MD, USA

192 Broad Institute, Cambridge, MA, USA

193 Univ. Bordeaux, Inserm, Bordeaux Population Health Research Center, UMR 1219, Bordeaux, France

194 Bordeaux University Hospital, Department of Neurology, Memory Clinic, Bordeaux, France

195 Neurovascular Research Laboratory. Vall d’Hebron Institut of Research, Neurology and Medicine Departments-Universitat Autònoma de Barcelona. Vall d’Hebrón Hospital, Barcelona, Spain

196 University Medicine Greifswald, Department of Internal Medicine B, Greifswald, Germany

197 DZHK, Greifswald, Germany

198 Robertson Center for Biostatistics, University of Glasgow, Glasgow, UK

199 Hero DMC Heart Institute, Dayanand Medical College & Hospital, Ludhiana, India

200 Atherosclerosis Research Unit, Department of Medicine Solna, Karolinska Institutet, Stockholm, Sweden

201 Karolinska Institutet, Stockholm, Sweden

202 Division of Emergency Medicine, and Department of Neurology, Washington University School of Medicine, St. Louis, MO, USA

203 Tohoku Medical Megabank Organization, Sendai, Japan

204 Department of Psychiatry, Washington University School of Medicine, St. Louis, MO, USA

205 Department of Public Health and Caring Sciences / Geriatrics, Uppsala University, Uppsala, Sweden

206 Epidemiology and Prevention Group, Center for Public Health Sciences, National Cancer Center, Tokyo, Japan

207 Department of Internal Medicine and the Center for Clinical and Translational Science, The Ohio State University, Columbus, OH, USA

208 Institute of Neuroscience and Physiology, the Sahlgrenska Academy at University of Gothenburg, Goteborg, Sweden

209 Department of Basic and Clinical Neurosciences, King’s College London, London, UK

210 Department of Health Care Administration and Management, Graduate School of Medical Sciences, Kyushu University, Japan

211 Department of Medicine and Clinical Science, Graduate School of Medical Sciences, Kyushu University, Japan

212 Landspitali National University Hospital, Departments of Neurology & Radiology, Reykjavik, Iceland

213 Department of Neurology, Heidelberg University Hospital, Germany

214 Department of Neurology, Erasmus University Medical Center

215 Hospital Universitari Mutua Terrassa, Terrassa (Barcelona), Spain

216 Albert Einstein College of Medicine, Montefiore Medical Center, New York, NY, USA

217 John Hunter Hospital, Hunter Medical Research Institute and University of Newcastle, Newcastle, NSW, Australia

218 Centre for Prevention of Stroke and Dementia, Nuffield Department of Clinical Neurosciences, University of Oxford, UK

219 Department of Medical Sciences, Uppsala University, Uppsala, Sweden

220 Genetic and Genomic Epidemiology Unit, Wellcome Trust Centre for Human Genetics, University of Oxford, Oxford, UK

221 The Wellcome Trust Centre for Human Genetics, Oxford, UK

222 Beth Israel Deaconess Medical Center, Boston, MA, USA

223 Wake Forest School of Medicine, Wake Forest, NC, USA

224 Department of Neurology, University of Pittsburgh, Pittsburgh, PA, USA

225 BioBank Japan, Laboratory of Clinical Sequencing, Department of Computational biology and medical Sciences, Graduate school of Frontier Sciences, The University of Tokyo, Tokyo, Japan

226 Neurovascular Research Laboratory, Vall d’Hebron Institut of Research, Neurology and Medicine Departments-Universitat Autònoma de Barcelona. Vall d’Hebrón Hospital, Barcelona, Spain

227 Department of Biostatistics, University of Liverpool, Liverpool, UK

228 Wellcome Trust Centre for Human Genetics, University of Oxford, Oxford, UK

229 Institute of Genetic Epidemiology, Helmholtz Zentrum München - German Research Center for Environmental Health, Neuherberg, Germany

230 Department of Medicine I, Ludwig-Maximilians-Universität, Munich, Germany

231 DZHK (German Centre for Cardiovascular Research), partner site Munich Heart Alliance, Munich, Germany

232 Department of Cerebrovascular Diseases, Fondazione IRCCS Istituto Neurologico “Carlo Besta”, Milano, Italy

233 Karolinska Institutet, MEB, Stockholm, Sweden

234 University of Tartu, Estonian Genome Center, Tartu, Estonia, Tartu, Estonia

235 Department of Clinical and Experimental Sciences, Neurology Clinic, University of Brescia, Italy

236 Translational Genomics Unit, Department of Oncology, IRCCS Istituto di Ricerche Farmacologiche Mario Negri, Milano, Italy

237 Department of Genetics, Microbiology and Statistics, University of Barcelona, Barcelona, Spain

238 Psychiatric Genetics Unit, Group of Psychiatry, Mental Health and Addictions, Vall d’Hebron Research Institute (VHIR), Universitat Autònoma de Barcelona, Biomedical Network Research Centre on Mental Health (CIBERSAM), Barcelona, Spain

239 Department of Neurology, IMIM-Hospital del Mar, and Universitat Autònoma de Barcelona, Spain

240 IMIM (Hospital del Mar Medical Research Institute), Barcelona, Spain

241 National Institute for Health Research Comprehensive Biomedical Research Centre, Guy’s & St. Thomas’ NHS Foundation Trust and King’s College London, London, UK

242 Division of Health and Social Care Research, King’s College London, London, UK

243 FIMM-Institute for Molecular Medicine Finland, Helsinki, Finland

244 THL-National Institute for Health and Welfare, Helsinki, Finland

245 Iwate Tohoku Medical Megabank Organization, Iwate Medical University, Iwate, Japan

246 BHF Glasgow Cardiovascular Research Centre, Faculty of Medicine, Glasgow, UK

247 deCODE Genetics/Amgen, Inc., Reykjavik, Iceland

248 Icelandic Heart Association, Reykjavik, Iceland

249 Institute of Biomedicine, the Sahlgrenska Academy at University of Gothenburg, Goteborg, Sweden

250 Department of Epidemiology, University of Maryland School of Medicine, Baltimore, MD, USA

251 Institute of Cardiovascular and Medical Sciences, Faculty of Medicine, University of Glasgow, Glasgow, UK

252 Chair of Genetic Epidemiology, IBE, Faculty of Medicine, LMU Munich, Germany

253 Division of Epidemiology and Prevention, Aichi Cancer Center Research Institute, Nagoya, Japan

254 Department of Epidemiology, Nagoya University Graduate School of Medicine, Nagoya, Japan

255 University Medicine Greifswald, Institute for Community Medicine, SHIP-KEF, Greifswald, Germany

256 Department of Neurology, Caen University Hospital, Caen, France

257 University of Caen Normandy, Caen, France

258 Department of Internal Medicine, Erasmus University Medical Center, Rotterdam, Netherlands

259 Landspitali University Hospital, Reykjavik, Iceland

260 Survey Research Center, University of Michigan, Ann Arbor, MI, USA

261 University of Virginia Department of Neurology, Charlottesville, VA, USA

## References

1. Rapsomaniki, E., Timmis, A., George, J., Pujades-Rodriguez, M., Shah, A.D., Denaxas, S., White, I.R., Caulfield, M.J., Deanfield, J.E., Smeeth, L., et al. (2014). Blood pressure and incidence of twelve cardiovascular diseases: lifetime risks, healthy life-years lost, and age-specific associations in 1.25 million people. Lancet 383, 1899–1911.

2. Ehret, G.B., Ferreira, T., Chasman, D.I., Jackson, A.U., Schmidt, E.M., Johnson, T., Thorleifsson, G., Luan, J., Donnelly, L.A., Kanoni, S., et al. (2016). The genetics of blood pressure regulation and its target organs from association studies in 342,415 individuals. Nat Genet 48, 1171–1184.

3. Ehret, G.B., Munroe, P.B., Rice, K.M., Bochud, M., Johnson, A.D., Chasman, D.I., Smith, A.V., Tobin, M.D., Verwoert, G.C., Hwang, S.J., et al. (2011). Genetic variants in novel pathways influence blood pressure and cardiovascular disease risk. Nature 478, 103–109.

4. Hoffmann, T.J., Theusch, E., Haldar, T., Ranatunga, D.K., Jorgenson, E., Medina, M.W., Kvale, M.N., Kwok, P.Y., Schaefer, C., Krauss, R.M., et al. (2018). A large electronic-health-record-based genome-wide study of serum lipids. Nat Genet 50, 401–413.

5. Warren, H.R., Evangelou, E., Cabrera, C.P., Gao, H., Ren, M., Mifsud, B., Ntalla, I., Surendran, P., Liu, C., Cook, J.P., et al. (2017). Genome-wide association analysis identifies novel blood pressure loci and offers biological insights into cardiovascular risk. Nat Genet.

6. Evangelou, E., Warren, H.R., Mosen-Ansorena, D., Mifsud, B., Pazoki, R., Gao, H., Ntritsos, G., Dimou, N., Cabrera, C.P., Karaman, I., et al. (2018). Genetic analysis of over 1 million people identifies 535 new loci associated with blood pressure traits. Nat Genet 50, 1412–1425.

7. Franceschini, N., Fox, E., Zhang, Z., Edwards, T.L., Nalls, M.A., Sung, Y.J., Tayo, B.O., Sun, Y.V., Gottesman, O., Adeyemo, A., et al. (2013). Genome-wide association analysis of blood-pressure traits in African-ancestry individuals reveals common associated genes in African and non-African populations. Am J Hum Genet 93, 545–554.

8. Liang, J., Le, T.H., Edwards, D.R.V., Tayo, B.O., Gaulton, K.J., Smith, J.A., Lu, Y., Jensen, R.A., Chen, G., Yanek, L.R., et al. (2017). Single-trait and multi-trait genome-wide association analyses identify novel loci for blood pressure in African-ancestry populations. PLoS Genet 13, e1006728.

9. Zhu, X., Feng, T., Tayo, B.O., Liang, J., Young, J.H., Franceschini, N., Smith, J.A., Yanek, L.R., Sun, Y.V., Edwards, T.L., et al. (2015). Meta-analysis of correlated traits via summary statistics from GWASs with an application in hypertension. Am J Hum Genet 96, 21–36.

10. Liu, C., Kraja, A.T., Smith, J.A., Brody, J.A., Franceschini, N., Bis, J.C., Rice, K., Morrison, A.C., Lu, Y., Weiss, S., et al. (2016). Meta-analysis identifies common and rare variants influencing blood pressure and overlapping with metabolic trait loci. Nat Genet 48, 1162–1170.

11. Surendran, P., Drenos, F., Young, R., Warren, H., Cook, J.P., Manning, A.K., Grarup, N., Sim, X., Barnes, D.R., Witkowska, K., et al. (2016). Trans-ancestry meta-analyses identify rare and common variants associated with blood pressure and hypertension. Nat Genet 48, 1151–1161.

12. Giri, A., Hellwege, J.N., Keaton, J.M., Park, J., Qiu, C., Warren, H.R., Torstenson, E.S., Kovesdy, C.P., Sun, Y.V., Wilson, O.D., et al. (2019). Trans-ethnic association study of blood pressure determinants in over 750,000 individuals. Nat Genet 51, 51–62.

13. Sung, Y.J., Winkler, T.W., de Las Fuentes, L., Bentley, A.R., Brown, M.R., Kraja, A.T., Schwander, K., Ntalla, I., Guo, X., Franceschini, N., et al. (2018). A Large-Scale Multi-ancestry Genome-wide Study Accounting for Smoking Behavior Identifies Multiple Significant Loci for Blood Pressure. Am J Hum Genet 102, 375–400.

14. Sung, Y.J., de Las Fuentes, L., Winkler, T.W., Chasman, D.I., Bentley, A.R., Kraja, A.T., Ntalla, I., Warren, H.R., Guo, X., Schwander, K., et al. (2019). A multi-ancestry genome-wide study incorporating gene-smoking interactions identifies multiple new loci for pulse pressure and mean arterial pressure. Hum Mol Genet.

15. Schillaci, G., and Pucci, G. (2010). The dynamic relationship between systolic and diastolic blood pressure: yet another marker of vascular aging? Hypertension research : official journal of the Japanese Society of Hypertension 33, 659–661.

16. Simino, J., Shi, G., Bis, J.C., Chasman, D.I., Ehret, G.B., Gu, X., Guo, X., Hwang, S.J., Sijbrands, E., Smith, A.V., et al. (2014). Gene-age interactions in blood pressure regulation: a large-scale investigation with the CHARGE, Global BPgen, and ICBP Consortia. Am J Hum Genet 95, 24–38.

17. Shi, G., Gu, C.C., Kraja, A.T., Arnett, D.K., Myers, R.H., Pankow, J.S., Hunt, S.C., and Rao, D.C. (2009). Genetic effect on blood pressure is modulated by age: the Hypertension Genetic Epidemiology Network Study. Hypertension 53, 35–41.

18. Dumitrescu, L., Brown-Gentry, K., Goodloe, R., Glenn, K., Yang, W., Kornegay, N., Pui, C.H., Relling, M.V., and Crawford, D.C. (2011). Evidence for age as a modifier of genetic associations for lipid levels. Ann Hum Genet 75, 589–597.

19. Lasky-Su, J., Lyon, H.N., Emilsson, V., Heid, I.M., Molony, C., Raby, B.A., Lazarus, R., Klanderman, B., Soto-Quiros, M.E., Avila, L., et al. (2008). On the replication of genetic associations: timing can be everything! Am J Hum Genet 82, 849–858.

20. Jiang, X., Holmes, C., and McVean, G. (2021). The impact of age on genetic risk for common diseases. PLoS Genet 17, e1009723.

21. Smith, G.D., and Ebrahim, S. (2003). ‘Mendelian randomization’: can genetic epidemiology contribute to understanding environmental determinants of disease? International journal of epidemiology 32, 1–22.

22. Bycroft, C., Freeman, C., Petkova, D., Band, G., Elliott, L.T., Sharp, K., Motyer, A., Vukcevic, D., Delaneau, O., O’Connell, J., et al. (2018). The UK Biobank resource with deep phenotyping and genomic data. Nature 562, 203–209.

23. McCarthy, S., Das, S., Kretzschmar, W., Delaneau, O., Wood, A.R., Teumer, A., Kang, H.M., Fuchsberger, C., Danecek, P., Sharp, K., et al. (2016). A reference panel of 64,976 haplotypes for genotype imputation. Nat Genet 48, 1279–1283.

24. Zhu, X., Li, X., Xu, R., and Wang, T. (2021). An iterative approach to detect pleiotropy and perform Mendelian Randomization analysis using GWAS summary statistics. Bioinformatics 37, 1390–1400.

25. Qi, G., and Chatterjee, N. (2019). Mendelian randomization analysis using mixture models for robust and efficient estimation of causal effects. Nat Commun 10, 1941.

26. Egger, M., Davey Smith, G., Schneider, M., and Minder, C. (1997). Bias in meta-analysis detected by a simple, graphical test. Bmj 315, 629–634.

27. Borenstein, M., Hedges, L., Higgins, J., Rothstein, H.. (2009). Generality of the basic inverse-variance method. In: Introduction to Meta-analysis. Chichester, UK: Wiley.

28. Burgess, S., Butterworth, A., and Thompson, S.G. (2013). Mendelian randomization analysis with multiple genetic variants using summarized data. Genet Epidemiol 37, 658–665.

29. Bulik-Sullivan, B.K., Loh, P.R., Finucane, H.K., Ripke, S., Yang, J., Schizophrenia Working Group of the Psychiatric Genomics, C., Patterson, N., Daly, M.J., Price, A.L., and Neale, B.M. (2015). LD Score regression distinguishes confounding from polygenicity in genome-wide association studies. Nat Genet 47, 291–295.

30. Pers, T.H., Karjalainen, J.M., Chan, Y., Westra, H.J., Wood, A.R., Yang, J., Lui, J.C., Vedantam, S., Gustafsson, S., Esko, T., et al. (2015). Biological interpretation of genome-wide association studies using predicted gene functions. Nature communications 6, 5890.

31. Cuellar-Partida, G., Lundberg, M., Kho, P.F., D’Urso, S., Gutierrez-Mondragon, L., and Hwang, L. (2019). Complex-Traits Genetics Virtual Lab: A community-driven web platform for post-GWAS analyses. bioRxiv.

32. Lee, S.H., Goddard, M.E., Wray, N.R., and Visscher, P.M. (2012). A better coefficient of determination for genetic profile analysis. Genet Epidemiol 36, 214–224.

33. Watanabe, K., Taskesen, E., van Bochoven, A., and Posthuma, D. (2017). Functional mapping and annotation of genetic associations with FUMA. Nature communications 8, 1826.

34. MacArthur, J., Bowler, E., Cerezo, M., Gil, L., Hall, P., Hastings, E., Junkins, H., McMahon, A., Milano, A., Morales, J., et al. (2017). The new NHGRI-EBI Catalog of published genome-wide association studies (GWAS Catalog). Nucleic acids research 45, D896–D901.

35. Inouye, M., Abraham, G., Nelson, C.P., Wood, A.M., Sweeting, M.J., Dudbridge, F., Lai, F.Y., Kaptoge, S., Brozynska, M., Wang, T., et al. (2018). Genomic Risk Prediction of Coronary Artery Disease in 480,000 Adults: Implications for Primary Prevention. Journal of the American College of Cardiology 72, 1883–1893.

36. Maier, R.M., Zhu, Z., Lee, S.H., Trzaskowski, M., Ruderfer, D.M., Stahl, E.A., Ripke, S., Wray, N.R., Yang, J., Visscher, P.M., et al. (2018). Improving genetic prediction by leveraging genetic correlations among human diseases and traits. Nature communications 9, 989.

37. Krapohl, E., Patel, H., Newhouse, S., Curtis, C.J., von Stumm, S., Dale, P.S., Zabaneh, D., Breen, G., O’Reilly, P.F., and Plomin, R. (2018). Multi-polygenic score approach to trait prediction. Mol Psychiatry 23, 1368–1374.

38. Richardson, T.G., Harrison, S., Hemani, G., and Davey Smith, G. (2019). An atlas of polygenic risk score associations to highlight putative causal relationships across the human phenome. eLife 8.

39. Chasman, D.I., Giulianini, F., Demler, O.V., and Udler, M.S. (2020). Pleiotropy-Based Decomposition of Genetic Risk Scores: Association and Interaction Analysis for Type 2 Diabetes and CAD. Am J Hum Genet 106, 646–658.

40. Udler, M.S., Kim, J., von Grotthuss, M., Bonas-Guarch, S., Cole, J.B., Chiou, J., Christopher, D.A.o.b.o.M., the, I., Boehnke, M., Laakso, M., et al. (2018). Type 2 diabetes genetic loci informed by multi-trait associations point to disease mechanisms and subtypes: A soft clustering analysis. PLoS Med 15, e1002654.

41. Nikpay, M., Goel, A., Won, H.H., Hall, L.M., Willenborg, C., Kanoni, S., Saleheen, D., Kyriakou, T., Nelson, C.P., Hopewell, J.C., et al. (2015). A comprehensive 1,000 Genomes-based genome-wide association meta-analysis of coronary artery disease. Nat Genet 47, 1121–1130.

42. Webb, T.R., Erdmann, J., Stirrups, K.E., Stitziel, N.O., Masca, N.G., Jansen, H., Kanoni, S., Nelson, C.P., Ferrario, P.G., Konig, I.R., et al. (2017). Systematic Evaluation of Pleiotropy Identifies 6 Further Loci Associated With Coronary Artery Disease. Journal of the American College of Cardiology 69, 823–836.

43. Malik, R., Chauhan, G., Traylor, M., Sargurupremraj, M., Okada, Y., Mishra, A., Rutten-Jacobs, L., Giese, A.K., van der Laan, S.W., Gretarsdottir, S., et al. (2018). Multiancestry genome-wide association study of 520,000 subjects identifies 32 loci associated with stroke and stroke subtypes. Nat Genet 50, 524–537.

44. Franklin, S.S. (2004). Pulse pressure as a risk factor. Clinical and experimental hypertension 26, 645–652.

45. Khan, S.S., Cooper, R., and Greenland, P. (2020). Do Polygenic Risk Scores Improve Patient Selection for Prevention of Coronary Artery Disease? JAMA 323, 614–615.

46. Elliott, J., Bodinier, B., Bond, T.A., Chadeau-Hyam, M., Evangelou, E., Moons, K.G.M., Dehghan, A., Muller, D.C., Elliott, P., and Tzoulaki, I. (2020). Predictive Accuracy of a Polygenic Risk Score-Enhanced Prediction Model vs a Clinical Risk Score for Coronary Artery Disease. JAMA 323, 636–645.

47. Mosley, J.D., Gupta, D.K., Tan, J., Yao, J., Wells, Q.S., Shaffer, C.M., Kundu, S., Robinson-Cohen, C., Psaty, B.M., Rich, S.S., et al. (2020). Predictive Accuracy of a Polygenic Risk Score Compared With a Clinical Risk Score for Incident Coronary Heart Disease. JAMA 323, 627–635.

48. Franklin, S.S. (1999). Ageing and hypertension: the assessment of blood pressure indices in predicting coronary heart disease. Journal of hypertension Supplement:official journal of the International Society of Hypertension 17, S29–36.

49. Demange, P.A., Malanchini, M., Mallard, T.T., Biroli, P., Cox, S.R., Grotzinger, A.D., Tucker-Drob, E.M., Abdellaoui, A., Arseneault, L., van Bergen, E., et al. (2021). Investigating the genetic architecture of noncognitive skills using GWAS-by-subtraction. Nat Genet 53, 35–44.

50. Boyle, E.A., Li, Y.I., and Pritchard, J.K. (2017). An Expanded View of Complex Traits: From Polygenic to Omnigenic. Cell 169, 1177–1186.

51. Liu, X., Li, Y.I., and Pritchard, J.K. (2019). Trans Effects on Gene Expression Can Drive Omnigenic Inheritance. Cell 177, 1022–1034 e1026.

52. Chakravarti, A., and Turner, T.N. (2016). Revealing rate-limiting steps in complex disease biology: The crucial importance of studying rare, extreme-phenotype families. BioEssays : news and reviews in molecular, cellular and developmental biology 38, 578–586.

